# MedMatch: a standardized medication order format for data science and natural language processing applications

**DOI:** 10.64898/2026.01.13.26343949

**Authors:** Kaitlin Blotske, Kelli Henry, Xingmeng Zhao, Tianle Li, Adeleine Tilley, Moriah Cargile, Brian Murray, Yanjun Gao, Susan E. Smith, Erin F. Barreto, Seth Bauer, Sunghwan Sohn, Tianming Liu, Tellen Bennett, Andrea Sikora

**Affiliations:** Department of Biomedical Informatics, University of Colorado Anschutz, Aurora, CO, USA; ATLAS Institute, University of Colorado Boulder, Boulder, CO, USA; University of Colorado Anschutz Medical Campus, Skaggs School of Pharmacy and Pharmaceutical Sciences. Department of Clinical Pharmacy, Aurora, CO, USA; University of Georgia College of Pharmacy, Department of Clinical and Administrative Pharmacy, Athens, GA, USA; Department of Pharmacy, Division of Nephrology and Hypertension, Department of Medicine, Mayo Clinic, Rochester, MN, USA; Cleveland Clinic, Cleveland, OH, USA; Department of Artificial Intelligence and Informatics Mayo Clinic, Rochester, MN, USA; Department of Computer Science, University of Georgia, Athens, GA, USA; University of Colorado School of Medicine, Department of Biomedical Informatics Aurora, CO; Department of Biomedical Informatics, University of Colorado Anschutz, Aurora, CO, USA, University of Georgia College of Pharmacy, Department of Clinical and Administrative Pharmacy, Augusta, GA, USA

**Keywords:** large language model, artificial intelligence, healthcare, medications, pharmacy

## Abstract

**Background:** Without automation, evaluation of medication management with large language models (LLMs) is largely limited to resource-intensive clinician evaluation. The purpose of this study was to develop a standardized medication structure (MedMatch) that supports automated evaluation.

**Methods:** Our goal was to create a medication order format with face validity to a clinician panel that maximizes clarity and safety. We used a multi-step process including (1) clinician survey of medication orders, (2) clinician survey of order appropriateness, (3) order sentence structure formation, (4) benchmark experiment development, and (5) GPT-5-chat testing across multiple experiments, which included assessment of ability to transform a medication order into MedMatch format and identify the route of an order sentence and ability to identify missing order components.

**Results:** First, lexical overlap analysis demonstrated pattern norms with specific components appearing most commonly in order of drug name, dose, unit, route, and frequency. Secondly, formal written orders consistently showed low omissions and high overlap and were deemed more appropriate as computer-generated orders. Third, MedMatch formats were created for oral and intravenous routes. Fourth, benchmark datasets and an abbreviation document were created. Fifth, GPT-5-Chat accuracy on MedMatch order standardization was highest in intravenous push (96.7%), oral liquid (86.1%), and intravenous intermittent (85%) categories. Overall performance was 77.6%. Identification of missing order components was improved with the MedMatch format (76.8% vs. 66.7%).

**Conclusions:** This is the first attempt to create and assess standardized medication order formatting to support benchmarking performance of LLMs. Limitations identified here demonstrate the need for medication-specific methodology.

## Background

Medication errors remain a common yet preventable source of morbidity and mortality.^1–3^ The advent of large language models (LLMs) may serve as a tool to improve medication safety, but without appropriate performance validation, this technology may also bring a new-age source of error, given important technical and practical limitations.^1–3^ Standardizing the communication structure of medication orders is an important foundational step for data science efforts that wish to safely participate in the medication use process.

Medications are a unique knowledge and reasoning domain that is not well captured by the larging training corpuses used for LLMs. A medication order is comprised of precisely constructed alphanumeric combinations that require specialized knowledge to interpret (e.g., carvedilol 25mg PO BID), yet it structurally falls somewhere between natural language and a rules-based computer coding language. A single medication order may be *written* in multiple formats (e.g., vancomycin 1 gram in 250 milliliters of 0.9% sodium chloride intravenous every 8 hours, vanc 1g TID, vancomycin 1000mg IV every 8 hours), but this same information may be communicated among healthcare professionals colloquially in a highly contextual fashion using accepted abbreviations and Latin terminology (e.g., “TID” for “three times daily” which is similar, but not equivalent, to “q8h” for every 8 hours) or shorthand and informal colloquialisms (e.g., “vanc one q eight”). Both of these styles are yet different than the highly formalized (yet institution specific) nature of electronic health record (EHR) formats (for which standard common data models have fallen short). Despite best practices for clarity of written medication orders from organizations, such as the Institute for Safe Medication Practices (ISMP) and The Joint Commission, widespread variability persists across different EHRs in how medication data are captured and represented.^4^

This combination of specialized syntax, documentation variability, and informal language complicates not only LLM performance, but also standardized evaluation. Traditionally, LLMs are evaluated using automated metrics focused on the quality of generated text, including fluency, coherence, accuracy, and relevance. Common metrics include perplexity, accuracy, F1-score, and Recall-Oriented Understudy for Gisting Evaluation (ROUGE) score. While these automated metrics increase evaluation throughput and reduce reliance on labor-intensive human review, they are poorly suited to medication-related tasks. For example, “cefepime 2,000mg every 8 hours” is clinically similar to “cefepime 2gm three times a day,” yet they yield low scores due to minimal elements overlap (i.e., just “cefepime” in this example). As a result, evaluation of LLM performance for medication-related tasks requires clinician annotation, which is both time- and resource-intensive. Development of an automated evaluation approach tailored to medication-related tasks has the potential to facilitate rapid and safe LLM development by automating performance grading. This is particularly salient as related benchmarking work for LLMs with medication tasks has shown a propensity for errors that require evaluation and correction prior to clinical use.^5–11^

Our goal was to develop a medication-specific order format intended for LLM use (MedMatch) that is in alignment with human clinician expectations. The ultimate intention of this standardized format is that it can facilitate both automated performance evaluation (PE) of future LLMs and a unifying language for AI tools involved with medication order generation.

## Methods

### Study design and oversight

The purpose of this study was to create and test a standardized medication order format intended to facilitate LLM applications. A structured process including two phases was used: Phase I – MedMatch Development and Phase II – MedMatch Testing (**Figure 1** summarizes the project workflow).

**Figure 1.**
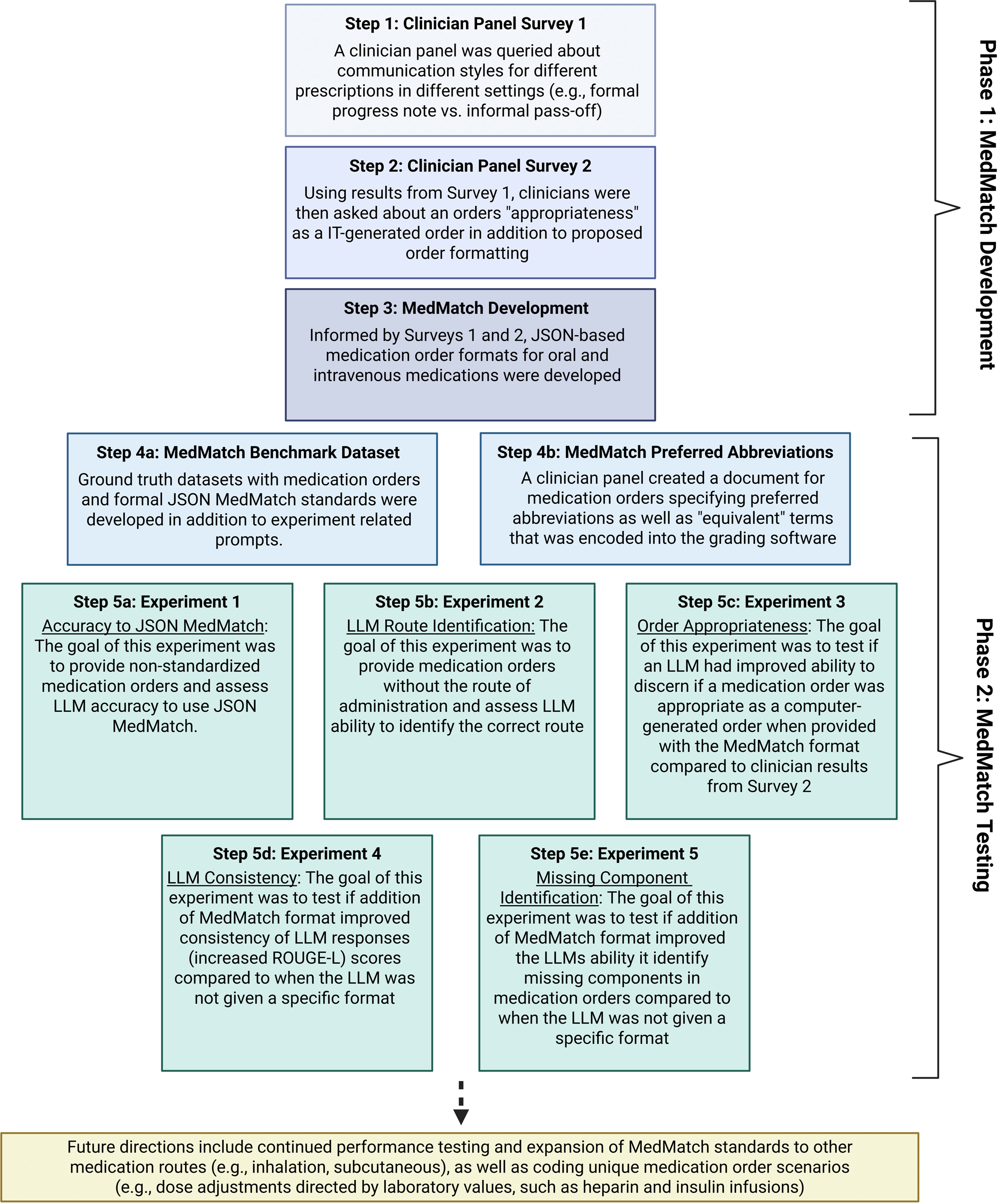
Methodological Flowchart. IT: information technology, JSON: JavaScript Object Notation, LLM: large language model, ROUGE: Recall-Oriented Understudy for Gisting Evaluation

This project was reviewed and approved by the University of Colorado Institutional Review Board (COMIRB #25-1631). All methods were performed in accordance with the ethical standards of the Helsinki Declaration of 1975.^12^ This evaluation followed the transparent reporting of a multivariable model for individual prognosis or diagnosis (TRIPOD–LLM) extension reporting frameworks, as applicable (**Supplemental Appendix**).^13^

### Phase I – MedMatch Development

Phase I consisted of three steps including two surveys (Steps 1 and 2) and MedMatch development (Step 3). Because of the variability in how human clinicians communicate medication order information (particularly in different contexts like formal progress notes vs. secure message systems among healthcare professionals), a panel of four board-certified clinical pharmacists was created to explore patterns in communication. Surveys were developed and administered via the Qualtrics^™^ platform (see **Supplemental Appendix)**. Results from Surveys 1 and 2 were employed by a panel of 5 clinicians and 2 computer scientists for an iterative development of the standardized medication order format using JavaScript Object Notation (JSON), which is a widely used language for software code: this format prompts the LLM to use a precise, structured data schema for its response, which can help for improving clarity and consistency (JSON prompts the LLM to behave more like a data processor, instead of a creative writer: this is also very true of how clinicians are trained to communicate medication orders because the risks of ambiguity that arise from natural language patterns are severe). The panel focused on identifying the information contained in a medication order that is required for safe and clear administration of different types of medications and determining an appropriate sequencing of information. Following descriptive analyses of these orders, the next step was the generation and discussion of proposed standardized medication order formats for medications with different formulations and routes of administration (**Table 1**).

**Table 1.** MedMatch medication order standards.

| Standard Type: | Standard in JSON Format: | Example: |
| --- | --- | --- |
| Oral solid dosage formulations | [drug name][numerical dose][abbreviated unit strength of dose][amount][formulation] by mouth [frequency] | Benzotropine 1 mg 1 tablet by mouth four times daily as needed |
| Oral liquid dosage formulations | [drug name][numerical dose][abbreviated unit strength of dose][numerical volume][abbreviated unit strength of volume] of the [concentration of formulation][formulation unit of measure][formulation] by mouth [frequency] | Diazepam 5 mg (5 mL) of the 1 mg/mL solution by mouth once daily |
| Intravenous intermittent dosing | [drug name][numerical dose][abbreviated unit strength of dose][amount of diluent volume][volume unit of measure] in [compatible diluent type] intravenously infused over [infusion time] [frequency] | Cefepime 2000 mg/100 mL in 0.9% sodium chloride intravenously infused over 30 minutes every 8 hours |
| Intravenous push dosing | [drug name][numerical dose][abbreviated unit strength of dose][amount of volume][volume unit of measure] of the [concentration of solution][concentration unit of measure][formulation] intravenous push [frequency] | Adenosine 6 mg (2 mL) of the 3 mg/mL vial solution intravenous push once |
| Continuous intravenous infusions | <u>Titratable:</u><br>[drug name][numerical dose][abbreviated unit strength of dose] [diluent volume][volume unit of measure] in [compatible diluent type] continuous intravenous infusion starting at [starting rate][unit of measure] titrated by [titration dose][titration unit of measure] [titration frequency] to achieve a goal of [titration goal]<br><u>Non-titratable:</u><br>[drug name][numerical dose][abbreviated unit strength of dose][diluent volume][volume unit of measure] in [compatible diluent type] continuous intravenous infusion at [starting rate][unit of measure] | Ketamine 500 mg/500 mL in 0.9% sodium chloride continuous intravenous infusion starting at 0.2 mg/kg/hr and titrated by 0.1 mg/kg/hr every 20 minutes to achieve a goal of RASS of -4 to -5<br>Vasopressin 20 units/100 mL in 0.9% sodium chloride continuous intravenous infusion at 0.03 units/min |

JSON: JavaScript Object Notation, kg: kilogram, mg: milligram, min: minute, mL: milliliter, RASS: Richmond Agitation and Sedation Scale

**Table 2.** Experiment 1 and 2: Accuracy to JSON MedMatch and LLM Route Identification (Steps 5a & 5b)

| Category | Experiment 1: Accuracy to JSON MedMatch |  |  |  | Experiment 2: LLM Route Identification |  |  |  |  |
| --- | --- | --- | --- | --- | --- | --- | --- | --- | --- |
|  | Correct fields, n/N | Field accuracy, % | All-fields-correct entries, n/N | Entry accuracy, % | n/run | Run 1, % | Run 2, % | Run 3, % | Mean, % |
| All medication categories | 7,974/8,280 | 96.3 | 698/900 | 77.6 | 300 | 96.7 | 96 | 96.3 | 96.3 |
| Oral solid | 1,200/1,260 | 95.2 | 123/180 | 68.3 | 60 | 100 | 100 | 100 | 100 |
| Oral liquid | 1,765/1,800 | 98.1 | 155/180 | 86.1 | 60 | 95 | 95 | 95 | 95 |
| Intravenous intermittent | 1,413/1,440 | 98.1 | 153/180 | 85.0 | 60 | 100 | 98.3 | 100 | 99.4 |
| Intravenous push | 1,605/1,620 | 99.1 | 174/180 | 96.7 | 60 | 90 | 88.3 | 88 | 88.9 |
| Intravenous continuous, titratable | 665/756 | 88.0 | 6/63 | 9.5 | 21 | 100 | 100 | 100 | 100 |
| Intravenous continuous, non-titratable | 1,326/1,404 | 94.4 | 87/117 | 74.4 | 39 | 97.4 | 97.4 | 97.4 | 97.4 |

Field accuracy = correct individual field comparisons / total field comparisons.
Entry accuracy = medication orders for which all evaluated fields were correct / total medication orders.
Route-identification accuracy = orders assigned to the correct route category / total orders.
Mean route accuracy is the arithmetic mean of the three run-specific percentages.

**Table 3.** Experiment 4: LLM Consistency (Step 5d)

| Consistency measure | Arm A: No format | Arm B: MedMatch format | Mean paired difference, B – A (95% CI) | Holm-adjusted p-value |
| --- | --- | --- | --- | --- |
| Modal exact repetitions out of 50 | 33.52 | 45.58 | 12.06 (10.47 to 13.66) | <0.001 |
| Number of distinct responses | 4.56 | 1.68 | –2.88 (–3.39 to –2.37) | <0.001 |
| Within-order mean pairwise ROUGE-L | 0.919 | 0.988 | 0.0694 (0.0596 to 0.0791) | <0.001 |
| Within-order median pairwise ROUGE-L | 0.937 | 0.991 | 0.0543 (0.0436 to 0.0649) | <0.001 |

**Table 4.** Experiment 5: Missing Component Identification (Step 5e)

| Components | Prompt A | Prompt B |
| --- | --- | --- |
| Oral solid<br>Numerical dose<br>Abbreviated unit of dose<br>Amount<br>Formulation<br>Route<br>Frequency | 29/30 (96.7%)<br>27/27 (100.0%)<br>0/24 (0.0%)<br>36/36 (100.0%)<br>30/33 (90.9%)<br>27/30 (90.0%) | 30/30 (100.0%)<br>27/27 (100.0%)<br>24/24 (100.0%)<br>35/36 (97.2%)<br>29/33 (87.9%)<br>27/30 (90.0%) |
| Oral liquid<br>Numerical dose<br>Abbreviated unit of dose<br>Numerical volume<br>Abbreviated unit of volume<br>Concentration of formulation<br>Formulation unit of measure<br>Formulation<br>Route<br>Frequency | 27/27 (100.0%)<br>24/24 (100.0%)<br>25/27 (92.6%)<br>19/24 (79.2%)<br>18/18 (100.0%)<br>0/12 (0.0%)<br>15/24 (62.5%)<br>12/12 (100.0%)<br>12/12 (100.0%) | 27/27 (100.0%)<br>24/24 (100.0%)<br>27/27 (100.0%)<br>22/24 (91.7%)<br>18/18 (100.0%)<br>0/12 (0.0%)<br>24/24 (100.0%)<br>12/12 (100.0%)<br>12/12 (100.0%) |
| Intravenous intermittent infusion<br>Numerical dose<br>Abbreviated unit of dose<br>Amount of diluent volume<br>Volume unit of measure<br>Compatible diluent type<br>Route<br>Infusion time<br>Frequency | 33/33 (100.0%)<br>27/27 (100.0%)<br>0/30 (0.0%)<br>14/18 (77.8%)<br>0/12 (0.0%)<br>12/12 (100.0%)<br>0/15 (0.0%)<br>33/33 (100.0%) | 29/33 (87.9%)<br>23/27 (85.2%)<br>0/30 (0.0%)<br>17/18 (94.4%)<br>11/12 (91.7%)<br>10/12 (83.3%)<br>9/15 (60.0%)<br>31/33 (93.9%) |
| Intravenous push<br>Numerical dose<br>Abbreviated unit of dose<br>Amount of volume<br>Volume unit of measure<br>Concentration of solution<br>Concentration unit of measure<br>Formulation<br>Route<br>Frequency | 18/18 (100.0%)<br>20/21 (95.2%)<br>15/18 (83.3%)<br>7/21 (33.3%)<br>18/18 (100.0%)<br>0/15 (0.0%)<br>0/12 (0.0%)<br>24/24 (100.0%)<br>33/33 (100.0%) | 18/18 (100.0%)<br>19/21 (90.5%)<br>15/18 (83.3%)<br>0/21 (0.0%)<br>0/18 (0.0%)<br>0/15 (0.0%)<br>0/12 (0.0%)<br>21/24 (87.5%)<br>33/33 (100.0%) |
| Continuous infusions (titratable)<br>Numerical dose<br>Abbreviated unit of dose<br>Diluent volume<br>Volume unit of measure<br>Compatible diluent type<br>Route<br>Starting rate<br>Unit of measure<br>Titration dose<br>Titration unit of measure<br>Titration frequency<br>Titration goal | 3/3 (100.0%)<br>—<br>0/3 (0.0%)<br>0/3 (0.0%)<br>—<br>—<br>—<br>—<br>0/9 (0.0%)<br>0/15 (0.0%)<br>0/15 (0.0%)<br>0/12 (0.0%) | 2/3 (66.7%)<br>—<br>0/3 (0.0%)<br>1/3 (33.3%)<br>—<br>—<br>—<br>—<br>9/9 (100.0%)<br>11/15 (73.3%)<br>7/15 (46.7%)<br>11/12 (91.7%) |
| Continuous infusions (non-titratable)<br>Numerical dose<br>Abbreviated unit of dose<br>Diluent volume | 12/12 (100.0%)<br>9/15 (60.0%)<br>0/12 (0.0%) | 10/12 (83.3%)<br>9/15 (60.0%)<br>0/12 (0.0%) |
| Volume unit of measure | 0/18 (0.0%) | 9/18 (50.0%) |
| Compatible diluent type | 0/15 (0.0%) | 8/15 (53.3%) |
| Route | 15/15 (100.0%) | 10/15 (66.7%) |
| Rate | 6/12 (50.0%) | 12/12 (100.0%) |
| Rate unit of measure | 0/21 (0.0%) | 18/21 (85.7%) |
| **Overall exact-match accuracy by run** |  |  |
| Run 1, all routes | 201/300 (67.0%) | 225/300 (75.0%) |
| Run 2, all routes | 201/300 (67.0%) | 234/300 (78.0%) |
| Run 3, all routes | 198/300 (66.0%) | 232/300 (77.3%) |
| **Across runs, all routes** | **600/900 (66.7%)** | **691/900 (76.8%)** |
| **Route-specific exact-match accuracy across runs** |  |  |
| Oral solid | 149/180 (82.8%) | 172/180 (95.6%) |
| Oral liquid | 152/180 (84.4%) | 166/180 (92.2%) |
| Intravenous intermittent | 119/180 (66.1%) | 130/180 (72.2%) |
| Intravenous push | 135/180 (75.0%) | 106/180 (58.9%) |
| Intravenous continuous | 45/180 (25.0%) | 117/180 (65.0%) |
| **All medication categories** | **600/900 (66.7%)** | **691/900 (76.8%)** |

Component-level and overall values report exact-match accuracy. Exact match required the response to identify exactly the withheld component, with no additional component or uncertainty statement; unparseable or uncanonicalizable responses were scored as incorrect. Route-specific values pool three runs (180 predictions per route and prompt), and total values pool 900 predictions per prompt.

### Phase II – MedMatch Testing

The testing phase consisted of 2 parts: MedMatch dataset development (Step 4a), identification of preferred abbreviations (Step 4b) and MedMatch testing (Step 5a-5e), including 5 distinct experiments (**Figure 1**).

#### Part 1. MedMatch Benchmark Dataset Development (Step 4a)

Three datasets were developed by three clinicians, consisting of 300 medication prompts each. All datasets included oral solid, oral liquid, intravenous intermittent infusion, intravenous push, and intravenous continuous infusion medications. The first dataset was developed to evaluate LLM performance in placing medication information into the MedMatch order standards, and the second dataset was developed to assess the LLMs’ ability to correctly determine medication route of administration based on contextual information void of specified routes. The third dataset was created with additional missing components (e.g., route, strength, dosing unit) to determine a single LLM’s performance to identify incomplete medication orders.

The first dataset included (1) a ground truth standardized medication order for LLM performance comparison, and (2) the prompt consisting of a medication order, in sentence format, for testing LLMs against the <u>MedMatch</u> formats. For example, a medication sentence format was presented as such: “The patient received amiodarone 150 mg prepared in 100 mL of D5W, administered intravenously and infused over 10 minutes once.” The second and third datasets were derived from the original by removing information about the route of administration from the standardized medication orders and removing additional important content from the medication orders (e.g., dose, frequency, infusion time).

#### Part 1. Preferred Abbreviation Document (Step 4b)

Additionally, a document was created that provided preferred MedMatch abbreviations as well as equivalent terms. This was used to improve accuracy of automated grading, by ensuring that interchangeable terms (e.g., mg and milligram) could be marked as correct, while also signifying what format is preferred for MedMatch. A clinician panel of 4 pharmacists reviewed this document and resolved any discrepancies through consensus voting. Abbreviations were avoided as the “preferred” term, with the exception of dosing units (i.e., mg is preferred over milligram or milligrams). The complete abbreviation document is provided in the **Supplementary Appendix**. For each field, the scorer checked the preferred-term and equivalent-expression families associated with both the ground-truth and predicted values. The field was considered correct only if the two values contained the same number of terms or phrases and their associated families overlapped at every corresponding position. The entire prediction was considered correct only if all fields were correct.

#### Part 2. MedMatch Testing (Steps 5a-5e)

A total of five experiments were conducted.

### Experiment 1: Accuracy to JSON MedMatch (Step 5a)

The first experiment used the clinician-curated dataset of 300 medication prompts to determine the accuracy of LLMs for arranging the medication sentence format prompts into the standardized medication order formats (**Table 1**). We evaluated GPT-5 Chat, given its impressive performance on other benchmarks.^5–7,14,15^ The medication order components were extracted in a single LLM call and then rendered into the MedMatch sentence format by deterministic Python code. The field accuracy was defined as the percentage of individual fields that matched the ground truth, whereas entry accuracy was defined as the percentage of medication orders for which all evaluated fields were correct. All prompts used for LLM evaluation are available in the **Supplemental Appendix**.

### Experiment 2: LLM Route Identification (Step 5b)

The second experiment was designed to probe the internal medication knowledge of the LLM. GPT-5 Chat was tested using a clinician-curated dataset of 300 medication orders in which route of administration information had been removed from standardized medication orders. The outcome of the second experiment was accuracy in categorizing the medications into defined route of administration categories: by mouth, intravenous push, intravenous intermittent infusion, and intravenous continuous infusion. The full dataset was run three times, yielding 900 predictions. Empty responses were retried up to three attempts and audited separately. LLM outputs underwent automated grading with clinician auditing.

### Experiment 3: Order Appropriateness (Step 5c)

The third experiment was designed to test the ability of an advanced LLM (GPT-5-Chat) to identify if a medication order is appropriate as a computer-generated order when provided with the MedMatch format compared to the model’s baseline knowledge without being provided the MedMatch format. This experiment used the dataset of 480 orders analyzed in Survey 2 and compared GPT-5-Chat to the human clinicians for analysis of verbal, brief written, and formal written orders. Percentage of medication orders deemed appropriate were reported for clinicians, LLM, clinicians with MedMatch, and LLM with MedMatch.

### Experiment 4: LLM Consistency (Step 5d)

The fourth experiment was designed to assess whether the MedMatch format improves the self-consistency of an LLM’s responses. GPT-5-Chat was tested using the 300 medication prompts in sentence format and was instructed to provide a complete medication order that would be appropriate as a computer-generated order on the medication administration record. Each medication was queried 100 times: 50 times with a prompt instructing the LLM only to standardize the medication order (Arm A), and 50 times with the same prompt plus the route-specific MedMatch format and a one-shot example (Arm B), for a total of 30,000 responses. The model was accessed through Azure at a temperature of 1.0. In both arms, the model was instructed to return the order as a single complete sentence without labels, JSON, bullets, or line breaks. Mean paired differences (Arm B minus Arm A) were reported with pointwise 95% confidence intervals. P-values were adjusted using the Holm method across the four consistency measures.

### Experiment 5: Missing Component Identification (Step 5e)

This experiment aimed to assess GPT-5-Chat’s ability to identify pertinent missing information from medication orders with and without the MedMatch standards. A dataset containing 300 medication orders with a variety of missing components (i.e., dose, strength, frequency, infusion time, etc.) was utilized for the experiment. Each medication order contained either 0 or 1 total omissions. The model was tested against two prompts. Prompt A asked the LLM to specify the missing component from each medication order and to specify uncertainty if unsure. Prompt B requested the same information, however, included the MedMatch Standards in JSON format with component definitions. A single example was provided in both prompts for one-shot promoting. Each medication order was run in triplicate for a total of 1800 outputs. LLM outputs underwent automated grading with clinician auditing.

### Statistical Analysis

Descriptive statistics, namely overall accuracy, were computed for each LLM response to the MedMatch testing for order sentence to JSON MedMatch format (Step 5a), LLM route identification (Step 5b), and missing medication order component identification (Step 5e). Accuracy was defined by number of entries where <u>all</u> required fields exactly match the corresponding ground truth standardized medication format (ignoring capitalization and additional spaces), divided by the total entries and multiplied by 100% for each LLM when prompted to place medication information into the MedMatch medication order standards. For the analysis of the LLM’s ability to appropriately categorize a medication order as appropriate as a computer-generated order (Step 5c), these results were reported as percentage and were compared with the human clinician assessment provided in Survey 2. For the LLM consistency experiment (Step 5d), a ROUGE-L and a self-consistency score were calculated for the responses without providing the MedMatch format, as well as when provided with the MedMatch format.

## Results

### Survey 1 (Step 1)

Lexical overlap results demonstrated pattern norms with specific components appearing most commonly in order of drug name, dose, unit, route, and frequency. The formal written orders consistently showed low omission rates and high overlap for all entities, compared to the verbal and brief written communications, which contained higher omissions of route and units. Detailed results are reported in the **Supplemental Appendix**.

### Survey 2 (Step 2)

The verbal communication and brief written communication medication orders had low rates of appropriateness. Within the formal written order group, oral medication prompts had higher rates of appropriateness labeling compared to intravenous medication prompts. Detailed results are reported in the **Supplemental Appendix**.

### MedMatch Standard Development (Step 3)

Clinician consensus resulted in six MedMatch standards: two oral medication order standards and four intravenous medication order standards, which are represented in **Table 1**. The two oral medication standards are delineated based on solid and liquid formulations, while the four intravenous medication standards were developed based on administration techniques of intermittent infusion, push dose, and continuous infusions (titratable and non-titratable). Panelist agreement for positioning of medication order content (in JSON format) was determined as: [drug name][dose][unit][route][frequency]. Additional content (e.g., volumes, concentrations, compatible diluents, titration doses) included in each MedMatch standard were deemed vital by clinician panelists for safety and completeness of medication orders.

### MedMatch Benchmark Dataset and Preferred Abbreviations (Steps 4a & 4b)

Two clinician-validated datasets were created, with 300 medication orders each. Additionally, an abbreviation document containing 136 abbreviations and equivalents was created and reported in the **Supplemental Appendix**. These documents were then used to improve automatic grading of experiments.

#### Experiment 1-Accuracy to JSON MedMatch (Step 5a)

Overall accuracy for each medication type (oral solid, intravenous push, etc.) is reported in **Table 2**. Each 300-order run yielded 2,760 field comparisons: 420 for oral solid orders (60 orders × 7 fields), 600 for oral liquid orders (60 × 10 fields), 480 for intravenous intermittent orders (60 × 8 fields), 540 for intravenous push orders (60 × 9 fields), and 720 for intravenous continuous-infusion orders (60 × 12 fields). The entire three run GPT-5 Chat achieved a 96.3% pooled field accuracy (7,974/8,280) and 77.6% pooled entry accuracy (698/900). Additional information about accuracy per order component (drug name, dose, dose unit, etc.) is reported in the **Supplementary Appendix**. GPT-5 Chat had the highest accuracy rate for intravenous push, oral liquid, and intravenous intermittent medications and had the lowest accuracy on titratable intravenous continuous infusions (9.5%) and oral solids (68.3%).

#### Experiment 2-LLM Route Identification (Step 5b)

Three full 300-order GPT-5 Chat runs yielded route identification accuracies of 96.67% (290/300), 96% (288/300), and 96.33% (289/300), with a mean of 96.33% (pooled 867/900 correct) (**Table 2**). Oral liquid accuracy was 95% in each run. Intravenous intermittent accuracy was 100%, 98.33%, and 100% across the three runs (mean 99.44%). Intravenous push had the lowest accuracy at 90%, 88.33%, and 88.33% (mean 88.89%). Intravenous continuous non-titratable accuracy was 97.44% across all three runs (**Table 2**).

#### Experiment 3: Order Appropriateness (Step 5c)

The LLM was more likely to label a medication order as “appropriate” when not provided with the MedMatch format, with brief written communication medication orders being labeled as appropriate more frequently by the LLM compared to the clinician evaluation of the same medication orders (**Figure 2**).

**Figure 2.**
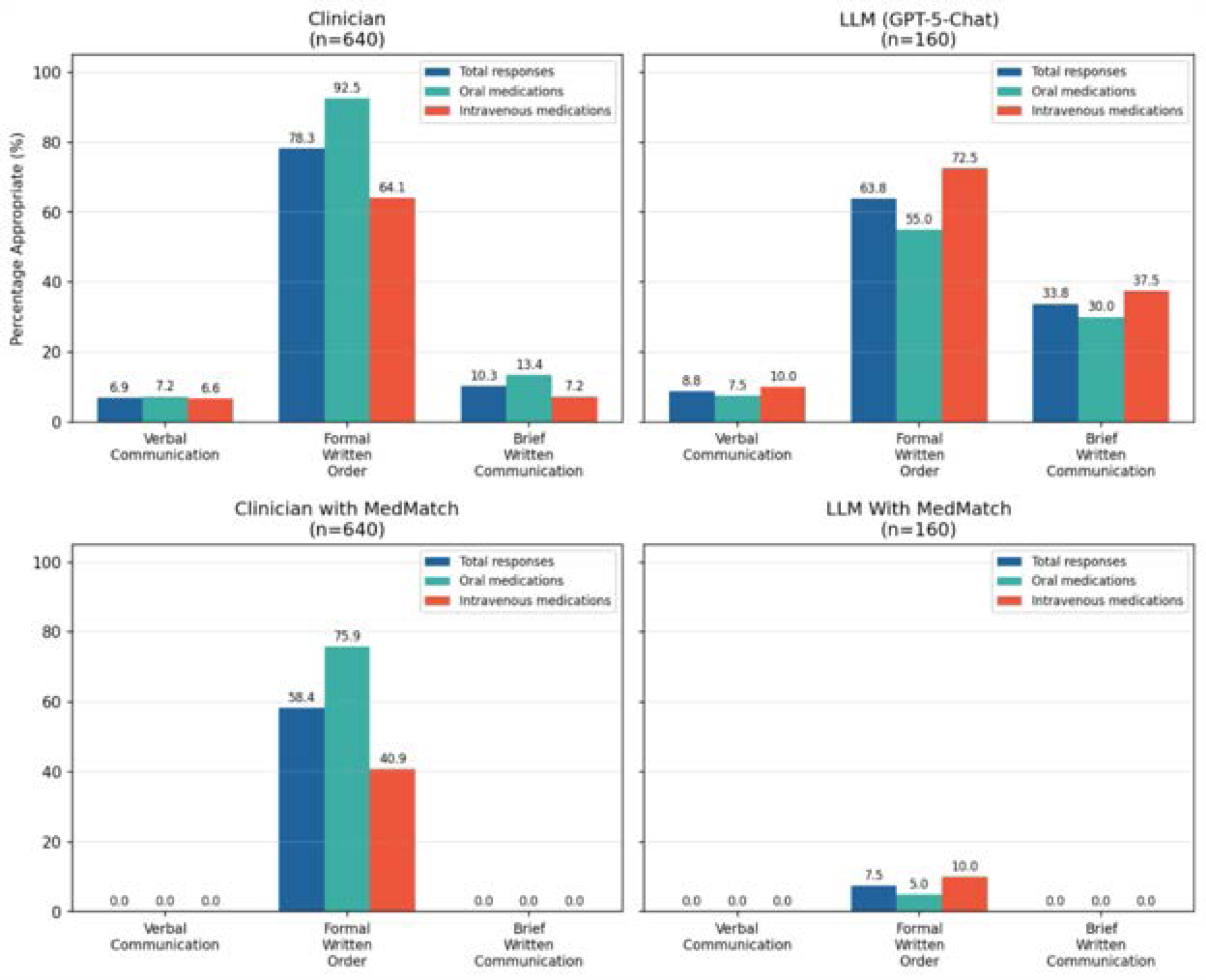
Experiment 3: Order Appropriateness. The figure depicts the percentage of orders that were deemed an appropriate computer-generated order by human clinicians, an LLM (GPT-5-Chat), clinician provided with MedMatch, and an LLM (GPT-5-Chat) provided with the MedMatch format that describes a complete order that would be considered an appropriate computer-generated order. Order types are divided both by the type of communication (formal written, brief written, and verbal) and the type of medication (intravenous or oral). A total of 640 clinician responses were evaluated for each order type (formal written, brief written, and verbal) compared to 160 LLM responses for each type with and without the MedMatch format provided in the prompt. LLM: large language model

In the subsequent survey, where clinicians and GPT-5-Chat were provided the MedMatch format alongside the same survey, results were similar with the formal written communication group remaining the highest for appropriateness as computer generated orders per clinician assessment. However, complete agreement occurred between clinicians with rated appropriateness dropping to 0% in both the verbal and brief written communication groups when compared to the MedMatch standards.

#### Experiment 4: LLM Consistency (Step 5d)

The MedMatch-format arm (Arm B) was more consistent than the no-format arm (Arm A) on all consistency measures (**Table 3**). The mean modal exact repetitions out of 50 was 45.58 with MedMatch format versus 33.52 without, a mean paired increase of 12.06 repetitions (95% CI, 10.47–13.66). The mean number of distinct responses per medication order was 1.68 versus 4.56. The averages across medication orders of within-order mean and median pairwise ROUGE-L were 0.988 versus 0.919 and 0.991 versus 0.937, respectively.

#### Experiment 5: Missing Component Identification (Step 5e)

Including the MedMatch standards in the prompt (Prompt B) improved missing-component identification accuracy compared to the baseline prompt (Prompt A) across all three replicate runs (**Table 4**). The overall accuracy was 76.8% with the MedMatch Prompt compared to 66.7% with the prompt that did not include the MedMatch format. GPT-5 had higher performance with the MedMatch prompt for all routes, with the exception of intravenous push (75% accuracy without MedMatch prompt compared to 58.9% accuracy with the MedMatch prompt).

## Discussion

This analysis is the first to develop a standardized medication order schema for LLM-generated medication orders, MedMatch. The intent of MedMatch is to facilitate safe communication of medication orders when using AI tools and to facilitate automated benchmarking of LLMs performance on medication-related tasks. MedMatch appeared to support clinician agreement regarding medication order communication.

Study findings suggest that formal orders exhibit the most stable linguistic conventions for all components of medication orders. Higher rates of variations between clinicians in verbal and brief communication styles highlight that communication reliability is influenced by both modality and individual practice patterns; however, formal written order communication was considered as the most widely accepted amongst clinicians as an automated response due to completeness of medication information. The addition of other medication components (volumes, diluents) during the MedMatch development phase emphasizes the complexities in safe medication ordering and administration. The six specific standards created here for each route (oral solid, oral liquid, IV push, IV intermittent, IV continuous titratable, IV continuous non-titratable) represent a clinical approach to the minimum required information needed for safe medication ordering.

MedMatch testing revealed variable performance. While some medication types exhibited reasonable performance (oral solid, intravenous push, and oral liquid), entry accuracy never exceeded 97% and for intravenous continuous titratable, entry accuracy was <10%. This may indicate that additional methodologies are needed to solidify MedMatch as a consistent option to improve automated evaluation of LLM results. Additionally, while route identification was impressive for some order types (oral solid and intravenous continuous), models exhibited variable accuracy for intravenous push and oral liquid medications. This may indicate an inability to identify how formulations and routes are interwoven, as noted in other analyses.^7^ Importantly, these limitations in present are likely solvable problems as the technology advances, but it does not eliminate the need for benchmarking of safety and ensuring product capabilities (this is somewhat akin to a safety checklist for a new vehicle including an item for seat belts and back-up cameras: the importance is in industry standardization and knowing what features affect safety).

Assessment of LLMs performance using the medication standards reflects the present limitations of LLMs for medication tasks and serves as a starting point for identifying future solutions. The unique nature of medication orders communication compared to natural language is a barrier to using LLMs. Medication orders include alphanumeric combinations of required information elements and have minimal margin for error without potentially lethal ramifications. The high-risk nature of medications requires standards to ensure both safe performance and clarity for use by clinicians. Although this clinician panel appeared to share similar unspoken rules, learned from training or professional recommendations (e.g., Institute for Safe Medication Practices), for structuring medication orders, no widely accepted standard for medication ordering currently exists.^4^ Beyond this, LLMs using progress notes or other natural language sources, which may omit critical safety information like route and frequency, observed in this study, may have knowledge gaps that are important for the LLM to recognize and find a safe way of clarifying (e.g., if a dose is omitted in a progress note, the LLM would need to recognize this and retrieve that dose before making a recommendation).

Emerging work probing the ability of LLMs to complete simple medication tasks has demonstrated substantial safety concerns.^8,9,16–18^ For example, LLMs currently lack the capability to distinguish a Pokémon from a drug, consistently identify drug-drug interactions, or perform tasks considered basic concepts developed during pharmacy school training.^6,7,10,11^ Using a framework similar to how clinicians are trained and licensed, entrustable professional activities (EPAs) have been proposed, which uses a benchmarking approach of standardized tasks and clinician-annotated datasets to ensure that a LLM can safely perform a given task.^17^ Automation of this grading process makes PE more efficient and may provide a baseline for understanding essential safety parameters needed for these technologies.

This study has several limitations. First, there exists the possibility of multiple reasonable means to communicate medication order information and that different approaches (e.g., modified Delphi) could yield different standards: there may be inherent benefits to other options that this study cannot rule out. Not all types of medication order are captured or validated within the provided standards (e.g., inhaled or subcutaneous routes). This work represents a pilot attempt at standardization, while recognizing the major complexities inherent to medication therapy. Future directions should include continued performance testing and expansion of MedMatch standards to other medication routes (e.g., inhalation, subcutaneous, intramuscular, ophthalmic) as well as coding unique medication order scenarios (e.g., dose adjustments directed by laboratory values, such as heparin and insulin infusions). These additional routes and scenarios make up a significant portion of medication complexity and have yet to be explored with the MedMatch standard. Additionally, limited models and testing scenarios were employed. While models were chosen for representativeness and cost constraints, ultimate deployment feasibility requires careful evaluation of parameter counts, context windows, or cost tradeoffs: the innovation presented here is that this approach could be used with any new models as a baseline safety check. Future directions could employ few-shot, chain-of-thought, and additional fine-tuning to improve performance. Lastly, these standards will ultimately require testing with more robust clinical datasets and clinician validation to be considered for full LLM performance automation.

## Conclusion

MedMatch serves as a potential standard for medication order information PE and LLM-generated medication recommendation outputs. This standard was developed through a structured approach to improve alignment between clinician and machine. The ability to grade LLM outputs automatically is a step towards medication safety in the era of AI but true avoidance of harm requires real-world evidence that accounts for unexpected human-machine interactions and other clinical considerations.

## Data Availability

Data is available upon reasonable request from the authors.

## TRIPOD-LLM Checklist

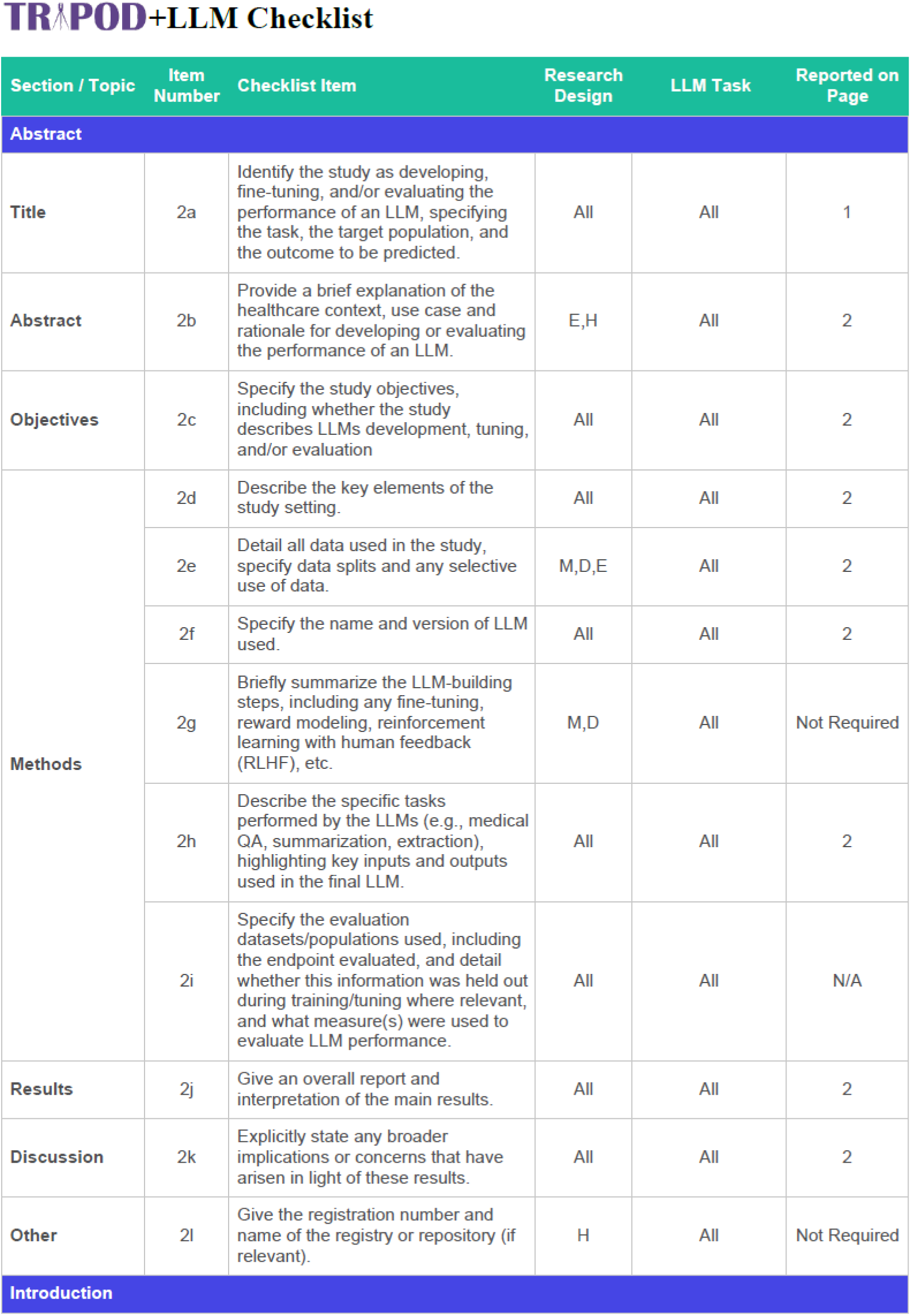
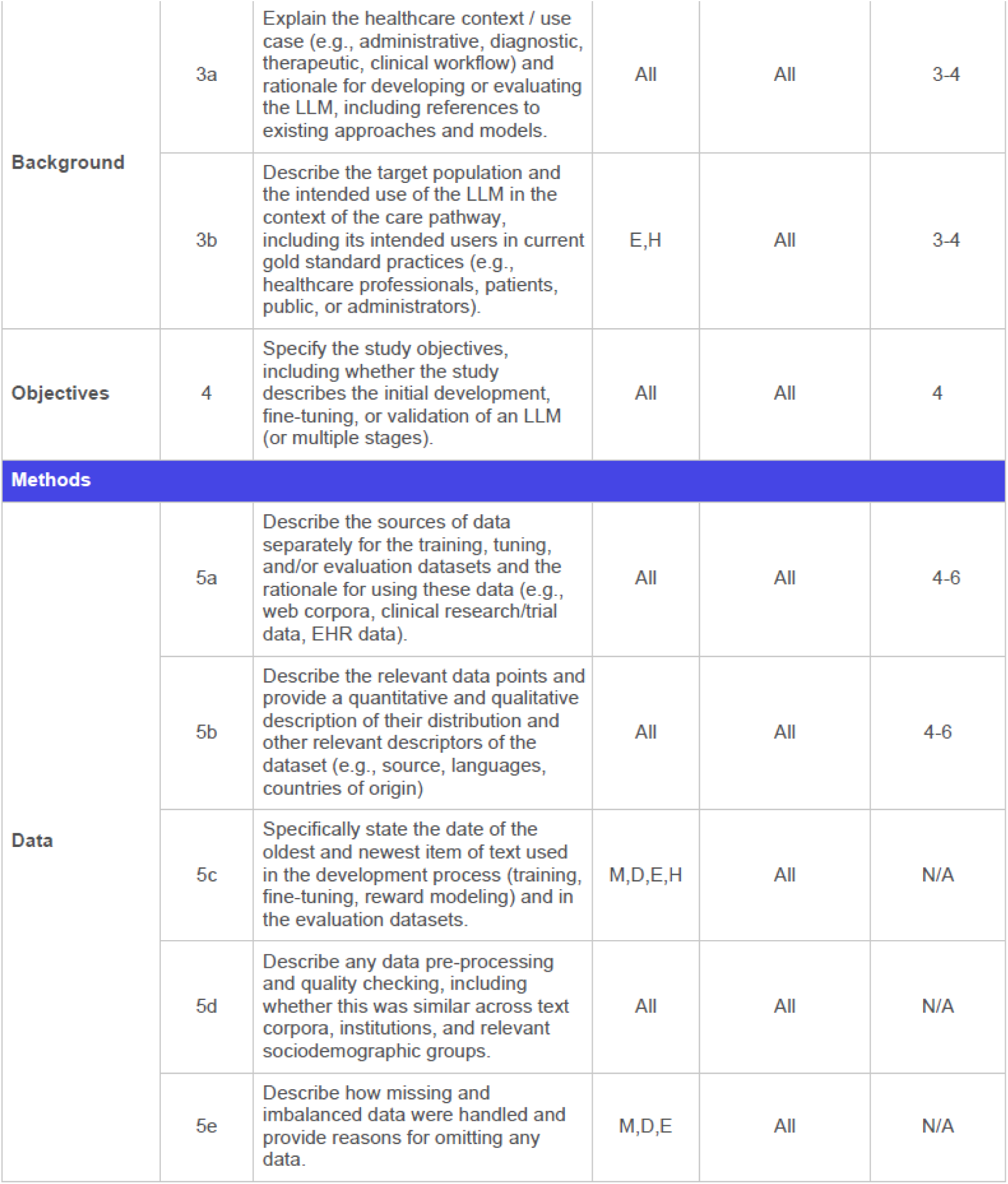
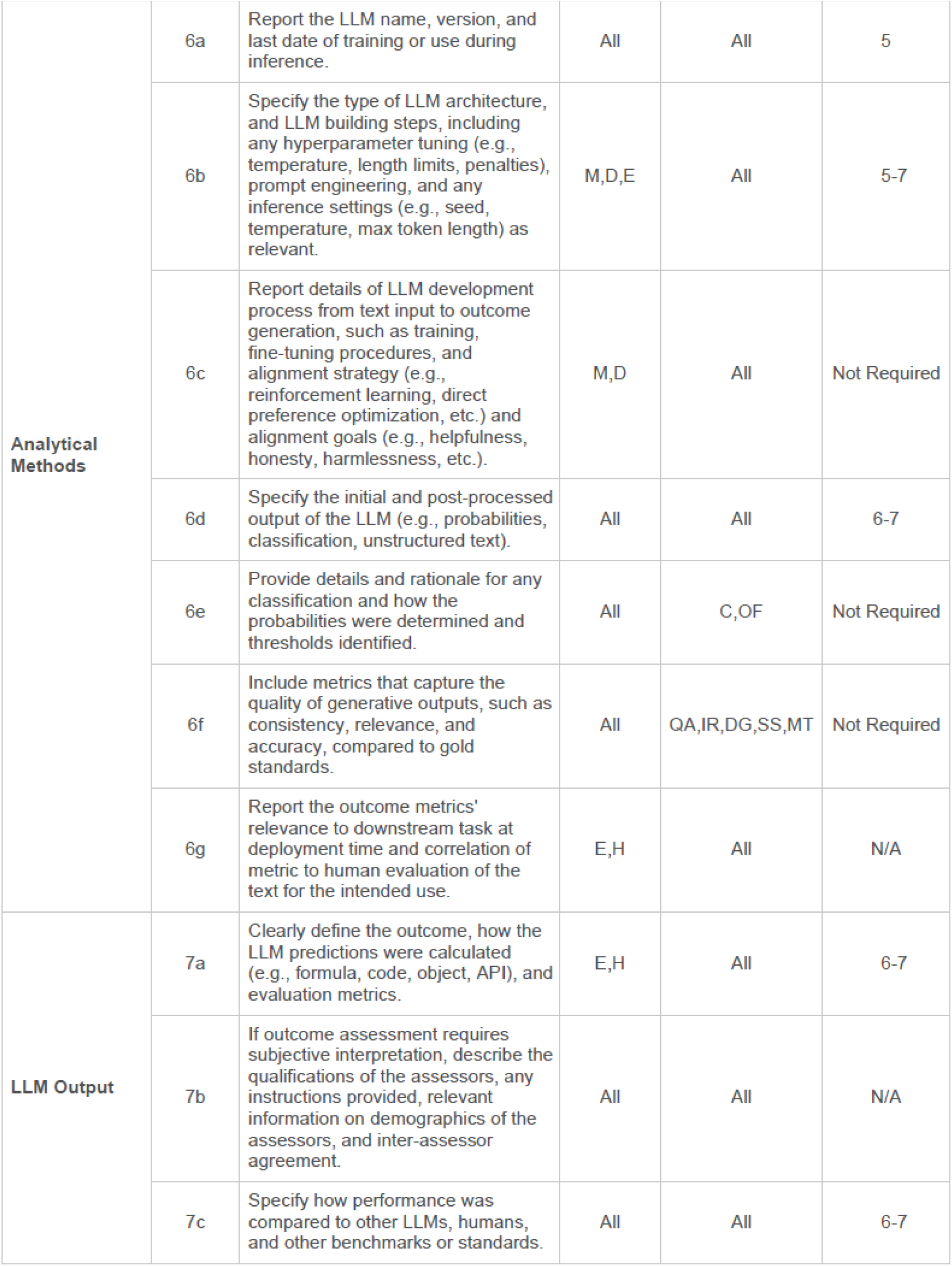
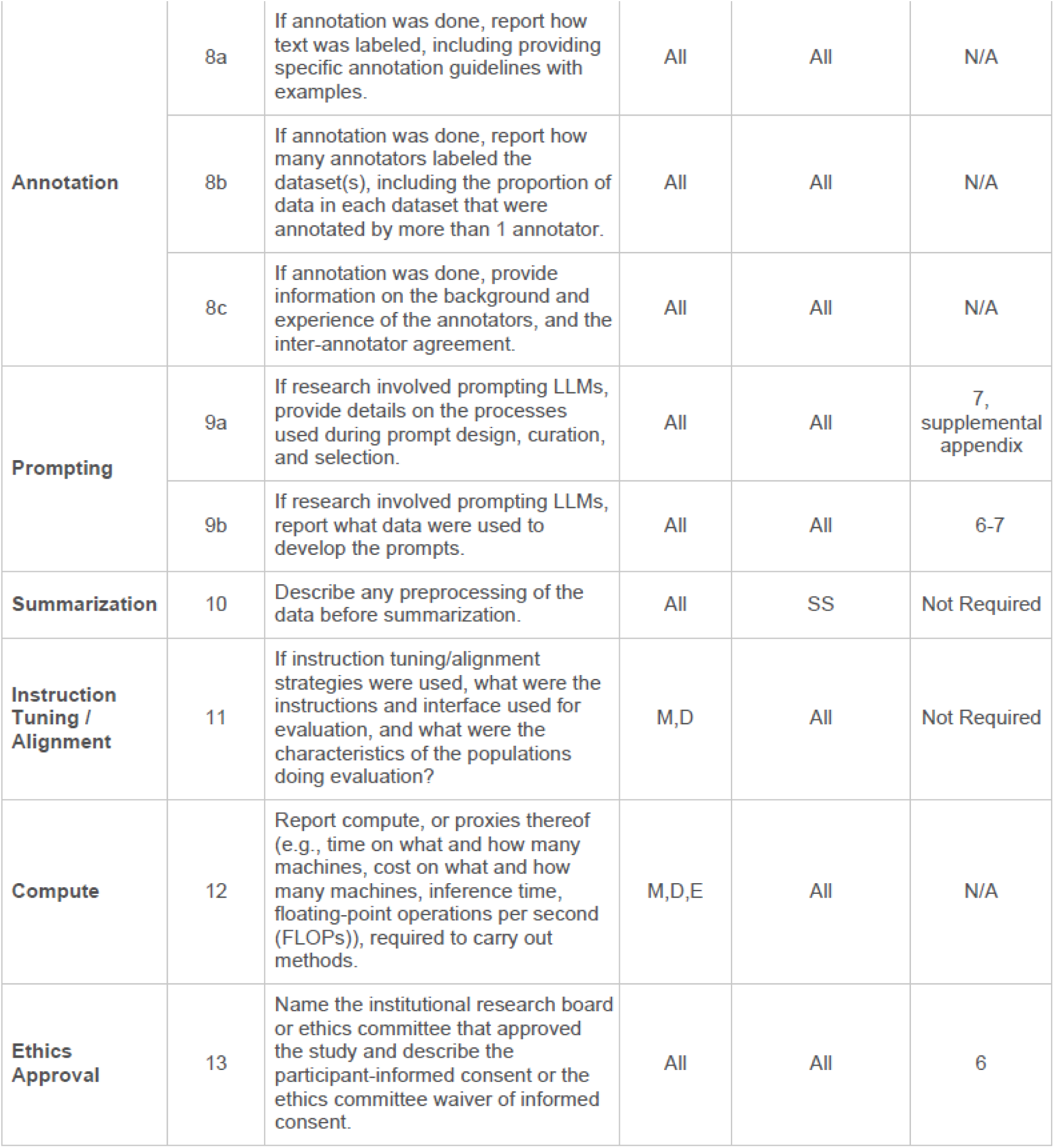
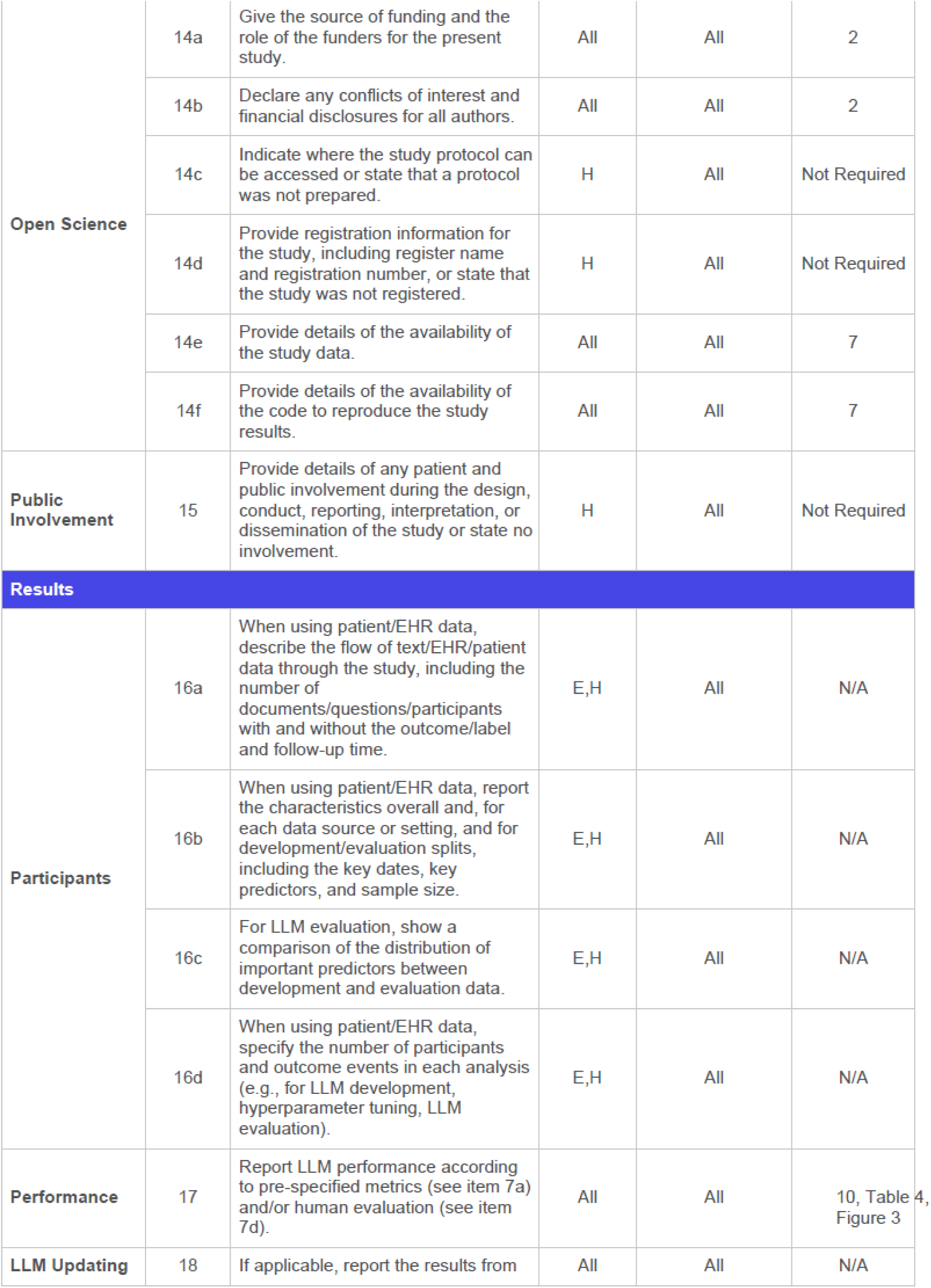
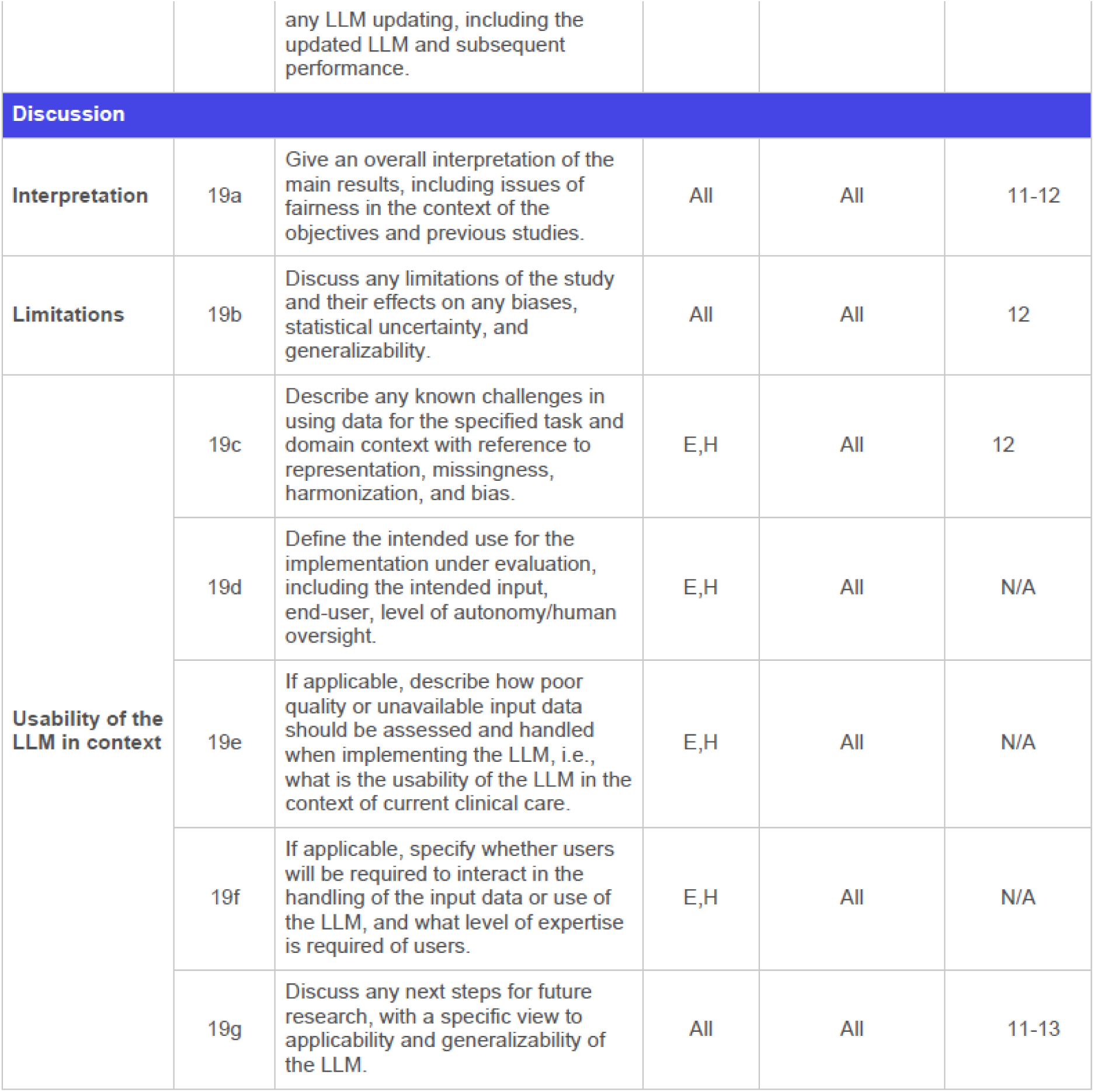

TRIPOD-LLM Checklist TRIPOD+LLM Checklist
| Section / Topic | Item Number | Checklist Item | Research Design | LLM Task | Reported on Page |
| --- | --- | --- | --- | --- | --- |
| **Abstract** |  |  |  |  |  |
| **Title** | 2a | Identify the study as developing, fine-tuning, and/or evaluating the performance of an LLM, specifying the task, the target population, and the outcome to be predicted. | All | All | 1 |
| **Abstract** | 2b | Provide a brief explanation of the healthcare context, use case and rationale for developing or evaluating the performance of an LLM. | E,H | All | 2 |
| **Objectives** | 2c | Specify the study objectives, including whether the study describes LLMs development, tuning, and/or evaluation | All | All | 2 |
| **Methods** | 2d | Describe the key elements of the study setting. | All | All | 2 |
|  | 2e | Detail all data used in the study, specify data splits and any selective use of data. | M,D,E | All | 2 |
|  | 2f | Specify the name and version of LLM used. | All | All | 2 |
|  | 2g | Briefly summarize the LLM-building steps, including any fine-tuning, reward modeling, reinforcement learning with human feedback (RLHF), etc. | M,D | All | Not Required |
|  | 2h | Describe the specific tasks performed by the LLMs (e.g., medical QA, summarization, extraction), highlighting key inputs and outputs used in the final LLM. | All | All | 2 |
|  | 2i | Specify the evaluation datasets/populations used, including the endpoint evaluated, and detail whether this information was held out during training/tuning where relevant, and what measure(s) were used to evaluate LLM performance. | All | All | N/A |
| **Results** | 2j | Give an overall report and interpretation of the main results. | All | All | 2 |
| **Discussion** | 2k | Explicitly state any broader implications or concerns that have arisen in light of these results. | All | All | 2 |
| **Other** | 2l | Give the registration number and name of the registry or repository (if relevant). | H | All | Not Required |
| **Introduction** |  |  |  |  |  |
| Background | 3a | Explain the healthcare context / use case (e.g., administrative, diagnostic, therapeutic, clinical workflow) and rationale for developing or evaluating the LLM, including references to existing approaches and models. | All | All | 3-4 |
|  | 3b | Describe the target population and the intended use of the LLM in the context of the care pathway, including its intended users in current gold standard practices (e.g., healthcare professionals, patients, public, or administrators). | E,H | All | 3-4 |
| Objectives | 4 | Specify the study objectives, including whether the study describes the initial development, fine-tuning, or validation of an LLM (or multiple stages). | All | All | 4 |
| Methods |  |  |  |  |  |
| Data | 5a | Describe the sources of data separately for the training, tuning, and/or evaluation datasets and the rationale for using these data (e.g., web corpora, clinical research/trial data, EHR data). | All | All | 4-6 |
|  | 5b | Describe the relevant data points and provide a quantitative and qualitative description of their distribution and other relevant descriptors of the dataset (e.g., source, languages, countries of origin) | All | All | 4-6 |
|  | 5c | Specifically state the date of the oldest and newest item of text used in the development process (training, fine-tuning, reward modeling) and in the evaluation datasets. | M,D,E,H | All | N/A |
|  | 5d | Describe any data pre-processing and quality checking, including whether this was similar across text corpora, institutions, and relevant sociodemographic groups. | All | All | N/A |
|  | 5e | Describe how missing and imbalanced data were handled and provide reasons for omitting any data. | M,D,E | All | N/A |
| Analytical Methods | 6a | Report the LLM name, version, and last date of training or use during inference. | All | All | 5 |
|  | 6b | Specify the type of LLM architecture, and LLM building steps, including any hyperparameter tuning (e.g., temperature, length limits, penalties), prompt engineering, and any inference settings (e.g., seed, temperature, max token length) as relevant. | M,D,E | All | 5-7 |
|  | 6c | Report details of LLM development process from text input to outcome generation, such as training, fine-tuning procedures, and alignment strategy (e.g., reinforcement learning, direct preference optimization, etc.) and alignment goals (e.g., helpfulness, honesty, harmlessness, etc.). | M,D | All | Not Required |
|  | 6d | Specify the initial and post-processed output of the LLM (e.g., probabilities, classification, unstructured text). | All | All | 6-7 |
|  | 6e | Provide details and rationale for any classification and how the probabilities were determined and thresholds identified. | All | C,OF | Not Required |
|  | 6f | Include metrics that capture the quality of generative outputs, such as consistency, relevance, and accuracy, compared to gold standards. | All | QA,IR,DG,SS,MT | Not Required |
|  | 6g | Report the outcome metrics' relevance to downstream task at deployment time and correlation of metric to human evaluation of the text for the intended use. | E,H | All | N/A |
| LLM Output | 7a | Clearly define the outcome, how the LLM predictions were calculated (e.g., formula, code, object, API), and evaluation metrics. | E,H | All | 6-7 |
|  | 7b | If outcome assessment requires subjective interpretation, describe the qualifications of the assessors, any instructions provided, relevant information on demographics of the assessors, and inter-assessor agreement. | All | All | N/A |
|  | 7c | Specify how performance was compared to other LLMs, humans, and other benchmarks or standards. | All | All | 6-7 |
| Annotation | 8a | If annotation was done, report how text was labeled, including providing specific annotation guidelines with examples. | All | All | N/A |
|  | 8b | If annotation was done, report how many annotators labeled the dataset(s), including the proportion of data in each dataset that were annotated by more than 1 annotator. | All | All | N/A |
|  | 8c | If annotation was done, provide information on the background and experience of the annotators, and the inter-annotator agreement. | All | All | N/A |
| Prompting | 9a | If research involved prompting LLMs, provide details on the processes used during prompt design, curation, and selection. | All | All | 7, supplemental appendix |
|  | 9b | If research involved prompting LLMs, report what data were used to develop the prompts. | All | All | 6-7 |
| Summarization | 10 | Describe any preprocessing of the data before summarization. | All | SS | Not Required |
| Instruction Tuning / Alignment | 11 | If instruction tuning/alignment strategies were used, what were the instructions and interface used for evaluation, and what were the characteristics of the populations doing evaluation? | M,D | All | Not Required |
| Compute | 12 | Report compute, or proxies thereof (e.g., time on what and how many machines, cost on what and how many machines, inference time, floating-point operations per second (FLOPs)), required to carry out methods. | M,D,E | All | N/A |
| Ethics Approval | 13 | Name the institutional research board or ethics committee that approved the study and describe the participant-informed consent or the ethics committee waiver of informed consent. | All | All | 6 |
| Open Science | 14a | Give the source of funding and the role of the funders for the present study. | All | All | 2 |
|  | 14b | Declare any conflicts of interest and financial disclosures for all authors. | All | All | 2 |
|  | 14c | Indicate where the study protocol can be accessed or state that a protocol was not prepared. | H | All | Not Required |
|  | 14d | Provide registration information for the study, including register name and registration number, or state that the study was not registered. | H | All | Not Required |
|  | 14e | Provide details of the availability of the study data. | All | All | 7 |
|  | 14f | Provide details of the availability of the code to reproduce the study results. | All | All | 7 |
| Public Involvement | 15 | Provide details of any patient and public involvement during the design, conduct, reporting, interpretation, or dissemination of the study or state no involvement. | H | All | Not Required |
| Results |  |  |  |  |  |
| Participants | 16a | When using patient/EHR data, describe the flow of text/EHR/patient data through the study, including the number of documents/questions/participants with and without the outcome/label and follow-up time. | E,H | All | N/A |
|  | 16b | When using patient/EHR data, report the characteristics overall and, for each data source or setting, and for development/evaluation splits, including the key dates, key predictors, and sample size. | E,H | All | N/A |
|  | 16c | For LLM evaluation, show a comparison of the distribution of important predictors between development and evaluation data. | E,H | All | N/A |
|  | 16d | When using patient/EHR data, specify the number of participants and outcome events in each analysis (e.g., for LLM development, hyperparameter tuning, LLM evaluation). | E,H | All | N/A |
| Performance | 17 | Report LLM performance according to pre-specified metrics (see item 7a) and/or human evaluation (see item 7d). | All | All | 10, Table 4, Figure 3 |
| LLM Updating | 18 | If applicable, report the results from any LLM updating, including the updated LLM and subsequent performance. | All | All | N/A |
| **Discussion** |  |  |  |  |  |
| **Interpretation** | 19a | Give an overall interpretation of the main results, including issues of fairness in the context of the objectives and previous studies. | All | All | 11-12 |
| **Limitations** | 19b | Discuss any limitations of the study and their effects on any biases, statistical uncertainty, and generalizability. | All | All | 12 |
| **Usability of the LLM in context** | 19c | Describe any known challenges in using data for the specified task and domain context with reference to representation, missingness, harmonization, and bias. | E,H | All | 12 |
|  | 19d | Define the intended use for the implementation under evaluation, including the intended input, end-user, level of autonomy/human oversight. | E,H | All | N/A |
|  | 19e | If applicable, describe how poor quality or unavailable input data should be assessed and handled when implementing the LLM, i.e., what is the usability of the LLM in the context of current clinical care. | E,H | All | N/A |
|  | 19f | If applicable, specify whether users will be required to interact in the handling of the input data or use of the LLM, and what level of expertise is required of users. | E,H | All | N/A |
|  | 19g | Discuss any next steps for future research, with a specific view to applicability and generalizability of the LLM. | All | All | 11-13 |

## Survey Structures Figure

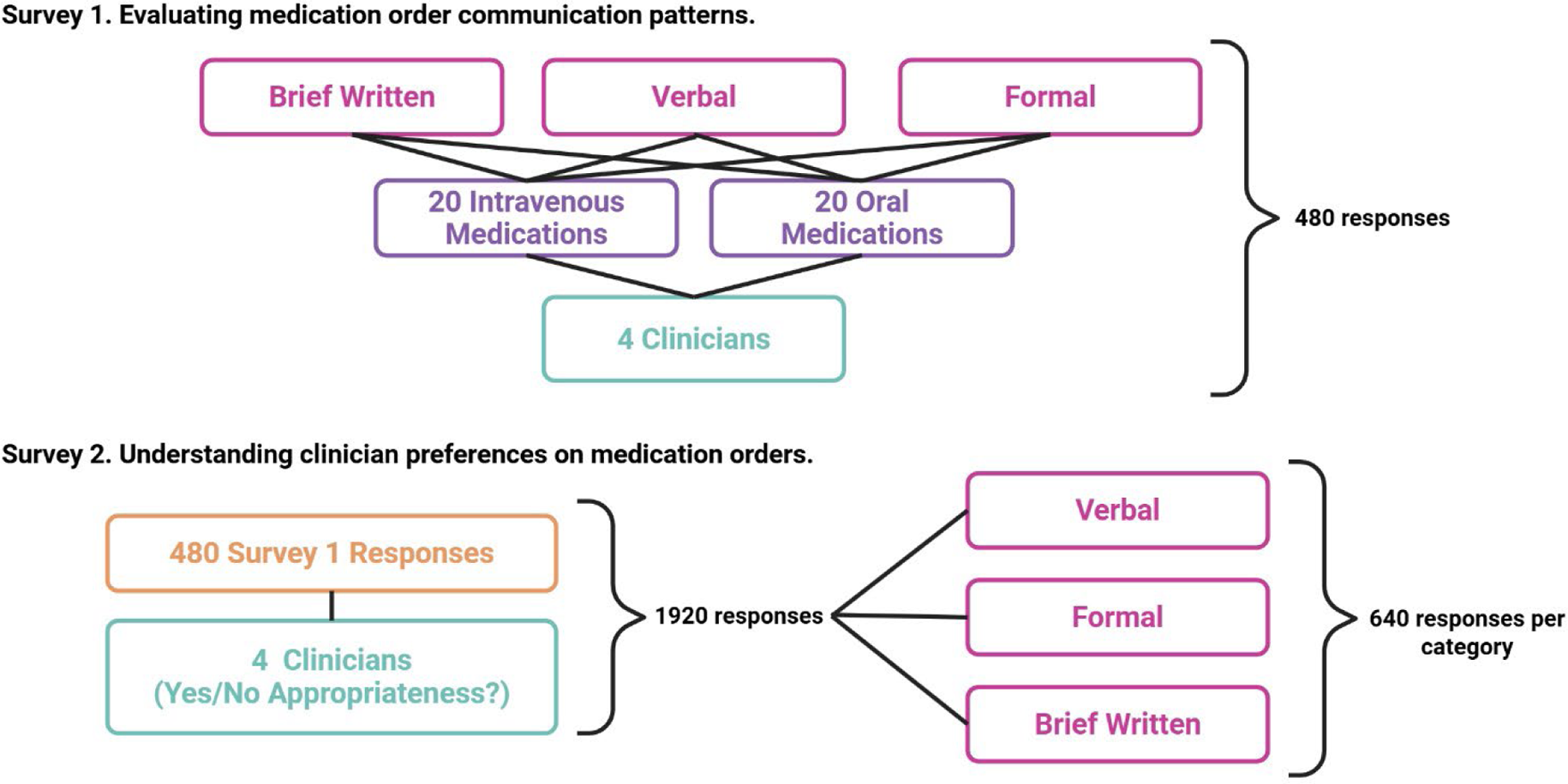

### Survey 1 Process

#### Survey to evaluate medication order communication patterns

The purpose of this survey was to generate baseline data assessing variability and the level of agreement for medication order communication across various contexts common to the healthcare setting. This survey consisted of 20 intravenous medication prompts and 20 oral medication prompts. For each prompt, respondents answered three questions: (1) How would you write this as a thorough and complete medication order? (2) How would you communicate this to another healthcare professional during rounds or hand-off? and (3) How would you convey this in a brief written message through a secure chat or other written communication with another clinician?

### Survey 1

#### Survey 1. Medication order baseline data generation for the detection of agreement levels via three communication scenarios

In this survey, you will be presented with brief prompts representing medication orders or clinical data sets, including both intravenous and non-intravenous medications. <u>For each prompt,</u> <u>please respond to the following three communication scenarios:</u>

1. **Formal Written Order** – How would you write this as a thorough and complete medication order?
2. **Verbal Communication** – How would you communicate this to another healthcare professional during rounds or hand-off?
3. **Brief Written Communication** – How would you convey this in a brief written message through a secure chat or other written communication with another clinician?

Please answer each question as if you are communicating with another clinician. There are 2 sections with 20 prompts in each section (40 prompts with 3 scenarios for each prompt) for a total of 120 entries.

In this first section, for each question you will be given a single prompt for an **oral** medication order that you should apply to the three communication scenarios to follow.

Prompts

Aspirin 81 mg daily

Atorvastatin 80mg daily

Carvedilol 25mg BID

Hydralazine 25mg TID

Acetaminophen 325mg q4h PRN headache

Warfarin 5mg MWF

Ibuprofen 800mg (as 200mg tablets) q8h PRN pain

Lisinopril 5mg qAM

Magnesium 400mgQID

Rimegepant 75mg QOD

Oxycodone 5mg q4h prn severe pain

Amlodipine 10mg every day

Cyclobenzaprine 15mg qHS

Amoxicillin 500mg BID

Sulfamethoxazole-trimethoprim (800-160) mg q12h

Prenatal vitamin daily

Metoprolol 25mg q6

Sertraline 25mg q day

Clopidogrel 75mg every day

Oxycodone 10mg q12h

In this final section, similar to before, you will be given a single prompt for an **intravenous** medication order or clinical data set that you should apply to the three communication scenarios to follow.

Prompts

Cefepime 1gm q24h

Cefepime 2gm q12h

Cefepime 2gm q8h

Norepinephrine 8mg/250mL at 5mcg/min

Vasopressin at 0.04units/min

Fentanyl at 50mcg/hr

Midazolam at 2mg/hr

0.9% sodium chloride at 100mL/hour

0.9% sodium chloride 1000mL over 30 minutes

Epinephrine 5mg/250mL @ 0.1mcg/kg/min

Propofol 1000mg/100mL starting at 5mcg/kg/min titrated to RASS 0 to −2

Dexmedetomidine 0.2mcg/kg/min

Ceftriaxone 2gm q24h

Cefazolin 2gm q8h

Meropenem 1gm BID

Dopamine 400mg/250mL @ 5mcg/min

Milrinone 40mg/200mL in D5W at 0.125mcg/kg/min

Hydrocortisone 50mg q6h

Hydromorphone 10mg/50mL @ 0.5mg/hr

D5W @ 200mL/hour

Survey 2 Process

#### Survey to understand clinician preference in terms of appropriateness of medication orders

The purpose of Survey 2 was to explore clinician preferences as it relates to medication order communication, particularly as generated by an information technology (IT) tool. For example, among healthcare professionals, “start the patient on baby aspirin” has inferred and colloquial meanings regarding aspirin dose, route, and frequency that are clear and unambiguous: “start the patient on aspirin 81mg by mouth once daily.” However, this shared meaning cannot be safely assumed beyond this context, and it is plausible that while a healthcare professional might use this shorthand, the preference from a safety perspective would be to overtly include dose, route, and frequency in the communication. The 480 responses generated in Survey 1 were subsequently used as evaluation prompts for Survey 2, which was distributed to the same clinician panel. In Survey 2, respondents were asked to assess whether each response would be appropriate if it were provided as a computer-generated output. Survey 2 was further composed into a two-step evaluation where clinicians were prompted with the above question, first without any proposed format and then with a proposed format for reference. Responses were recorded in a yes/no format.

### Survey 2

In this survey, you will be presented with the clinician medication orders responses generated from four critical care pharmacists in **Survey 1** for both intravenous and non-intravenous medications. <u>For each medication order, please judge responses using yes/no to the following</u> <u>question:</u>

### Would these clinical responses provided by clinical pharmacists be appropriate if provided as a “computer-generated” output?

Each clinical pharmacist provided 120 responses in **Survey 1**, resulting in a total of 480 responses that required an appropriateness assessment.

## Survey Additional Methods

For each survey, the extracted medication components (drug name, dose, unit, route, and frequency) were analyzed using word-level Jaccard similarity.^9^ All strings were lowercased and split on whitespace, and similarity was computed as the size of the intersection of word sets divided by the size of their union. To provide an overall indication of how much responses varied for each component, we calculated a variation score defined as 1 − *avg_jjaccard_*, where *avg_jjaccard_* is the mean pairwise similarity across all responses with majority answer. To compute Jaccard similarity, each string was lowercased and tokenized into words using whitespace splitting. The similarity score was then calculated as the size of the intersection of the resulting word sets divided by the size of their union. This offers a simple and interpretable way to capture how much the responses differ in wording.

For each question, agreement was analyzed at both the response level and the component level. At the component level, medication components (drug name, dose, unit, route, frequency) were extracted from each respondent’s full response text using GPT-4o-mini (temperature=0.1) with specialized prompts designed for each component type. This LLM-assisted extraction ensured accurate parsing of medication information from natural language responses. For each component, the majority value was identified as the most frequent non-empty extracted value across all respondents, ignoring empty strings and normalized missing values (e.g., “none”, “N/A”). Jaccard similarity was then computed between each respondent’s extracted component value and the majority value. The Jaccard similarity measures word-level overlap:

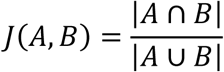

where *A* and *B* are word sets derived from the two component values. Text preprocessing involved converting to lowercase and tokenizing on whitespace boundaries, preserving all words including stop words to maintain semantic fidelity. This yields a continuous similarity score *J* ∈ [0,1], where *J* = 1 indicates identical word sets and *J* = 0 indicates no shared words. The Jaccard similarity provides a fine-grained measure of how similar each respondent’s component value is to the consensus, revealing both the degree of agreement and where responses deviate from the majority.

### Measuring Overlap

To quantify agreement between responses, overlap was measured at multiple levels using complementary approaches. While Jaccard similarity provides continuous similarity scores (

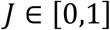

), both overlap percentage and pairwise overlap analysis use binary classification (overlap vs. no overlap) to assess agreement. These measures operate at different granularities: Jaccard similarity provides fine-grained continuous similarity scores between individual responses and the majority, overlap percentage uses a binary threshold (

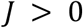

) to indicate the breadth of agreement (how many responses share content), and pairwise analysis uses binary classification to reveal overall consistency patterns across all respondent pairs.

### Overlap Percentage

To measure how many responses share any content with the consensus, an overlap percentage based on Jaccard similarity was computed. A response is considered to overlap with the majority if it shares at least one word with the majority value (i.e., its Jaccard similarity is greater than zero). The overlap percentage is then calculated as the proportion of responses that meet this criterion out of all responses. This produces a binary classification, responses either overlap with the majority or they do not, which complements the continuous Jaccard similarity scores by capturing the breadth of agreement. Thus, it indicates how many respondents align with the consensus at a minimal level, regardless of how similar their responses are beyond sharing any common words.

## Survey 1 Results

In the formal written medication order, we observed high completeness and consistency across all components with low omission rates, compared to verbal and brief written communication. The vocabulary used in formal orders is highly standardized, reflected by a high lexical overlap across panelists. Analysis of ordering patterns between the four clinicians further revealed strong structural regularities: drug names are positioned first in most orders, doses and units typically appear in the second and third position, route placement exhibits notably in the fourth position, and frequency most commonly in the fifth position for formal written orders (**Figure 1**). Ordering patterns in verbal communication show greater variability and context-driven speech patterns rather than the fixed structure observed in written orders. The brief written and verbal communication styles showed higher rates of omission of components as well as reduced overlap percentages in comparison to the formal written communication style.

**Figure 1.**
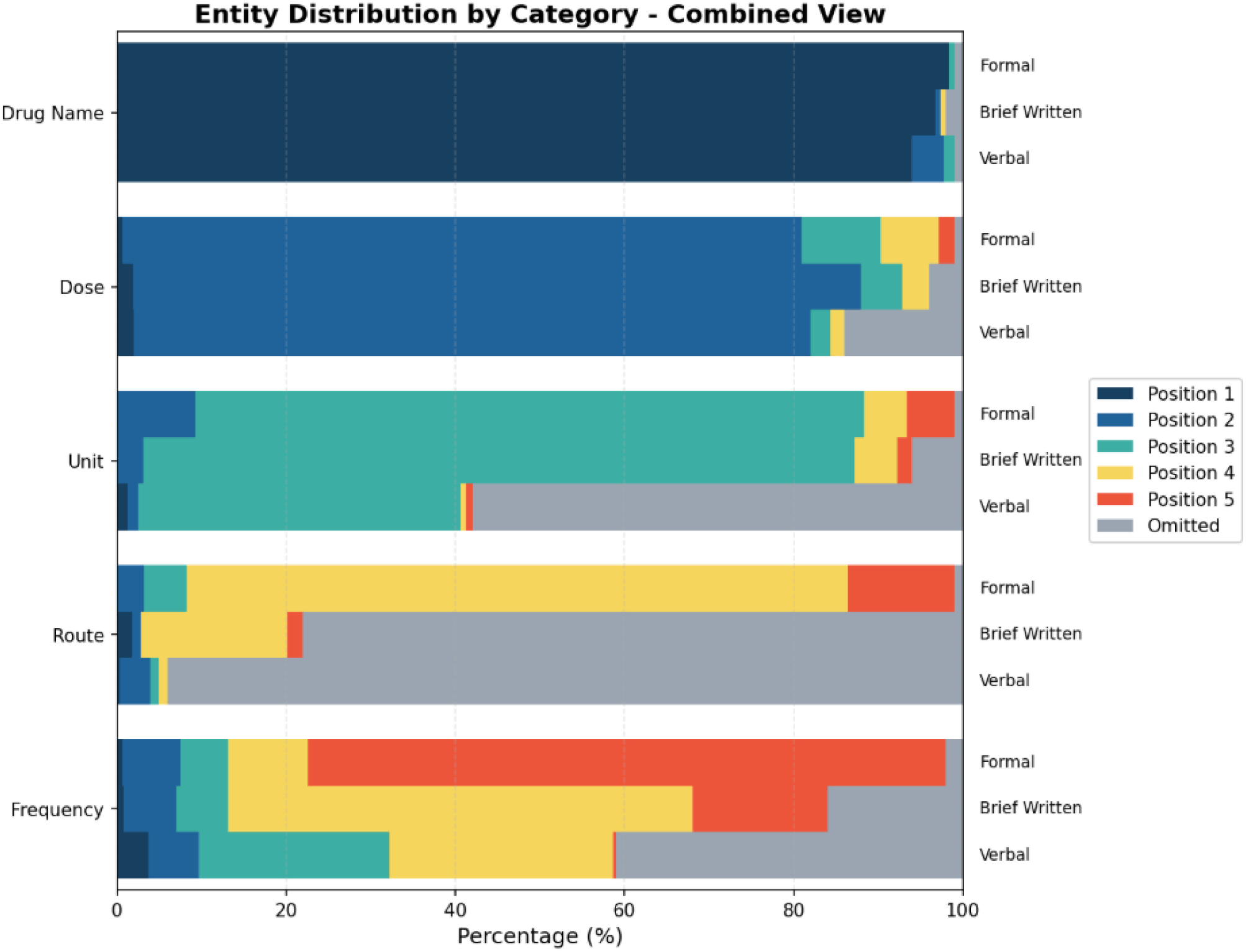
Survey 1 percentage of times each component is predicted in positions 1–5 for the initial clinician medication order survey results for formal written, brief written, and verbal communication styles. This figure shows how often each medication component (drug name, dose, unit, route, frequency) is placed in positions 1 through 5 or is omitted for each formal written, brief written and verbal communication styles from the initial survey completed by four clinician pharmacists. The colors in each entity row represent the percentage of predictions for each position, starting with position 1 (first mention) and ending with position 5 (last mention). Higher percentages in each color means the corresponding medication component is more likely to appear at that position per clinician communications.

**Table 1.** Survey 1 omission and overlap performance of clinician medication order survey.

| Question Type | Entity | Average Omission Percentage | Overlap Percentage | Clinician #1 Omission Percentage | Clinician #2 Omission Percentage | Clinician #3 Omission Percentage | Clinician #4 Omission Percentage |
| --- | --- | --- | --- | --- | --- | --- | --- |
| Formal (n=160) | Drug Name | 1 | 98 | 0 | 0 | 0 | 0 |
|  | Dose | 1 | 84 | 0 | 3 | 0 | 0 |
|  | Unit | 1 | 88 | 0 | 3 | 0 | 0 |
|  | Route | 1 | 95 | 3 | 3 | 0 | 0 |
|  | Frequency | 2 | 83 | 3 | 3 | 0 | 3 |
| Verbal<br>(n=160) | Drug Name | 1 | 66 | 0 | 0 | 3 | 0 |
|  | Dose | 14 | 89 | 30 | 8 | 5 | 13 |
|  | Unit | 58 | 56 | 80 | 90 | 3 | 58 |
|  | Route | 94 | 38 | 100 | 95 | 88 | 93 |
|  | Frequency | 41 | 36 | 60 | 48 | 3 | 55 |
| Brief<br>(n=160) | Drug Name | 2 | 83 | 0 | 3 | 3 | 3 |
|  | Dose | 4 | 90 | 5 | 5 | 5 | 3 |
|  | Unit | 6 | 73 | 8 | 10 | 5 | 3 |
|  | Route | 78 | 57 | 95 | 95 | 93 | 28 |
|  | Frequency | 16 | 63 | 33 | 10 | 13 | 8 |

This table summarizes the omission and overlap percentages for each Question Type (formal, verbal, brief) and Entity (drug name, dose, unit, route, frequency). “Omission%” and “Overlap%” represent the overall model performance, while the columns for Clinician #1-4 show the omission percentage contributed by each individual clinician. outputs. **Omission Percentage:** The percentage of times each medication component was completely missing from responses. **Overlap Percentage:** Word-level consistency between different respondents' answers for the same medication. Higher omission values indicate more missing information, whereas higher overlap values reflect better agreement with the reference.

**Table 2.**
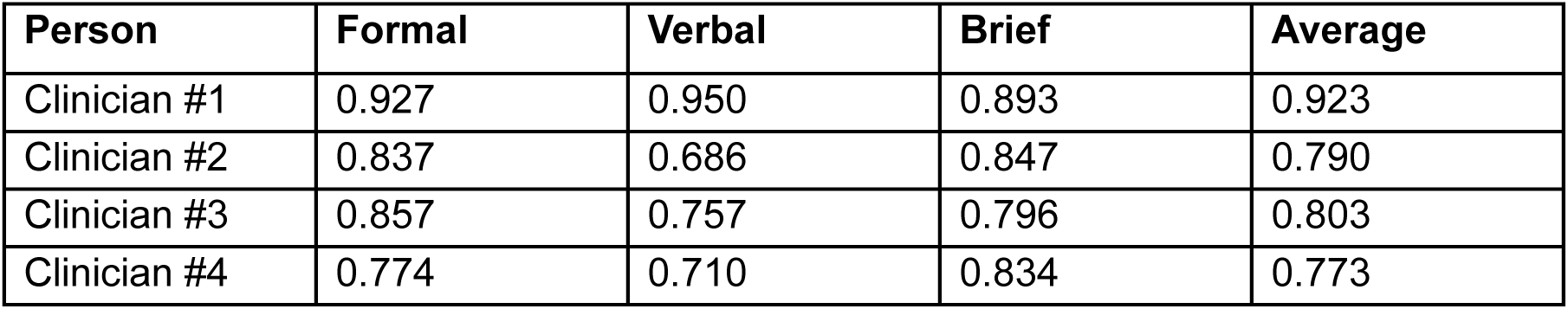
Survey 1 JACCARD similarity by individual and communication style for survey 1.

**Table 3.** Survey 1 Jaccard similarity assessment by component, communication type, and clinician.

| Variable | Average Jaccard Similarity |
| --- | --- |
| **Similarity for each component (across all communication styles)** |  |
| Drug Name | 0.880 |
| Dose | 0.897 |
| Unit | 0.778 |
| Route | 0.812 |
| Frequency | 0.706 |
| **Similarity by communication type (across all components)** |  |
| Formal | 0.847 |
| Verbal | 0.771 |
| Brief | 0.839 |
| **Similarity by clinician (across all components)** |  |
| Clinician #1 | 0.925 |
| Clinician #2 | 0.787 |
| Clinician #3 | 0.804 |
| Clinician #4 | 0.773 |

This table summarizes the average Jaccard similarity scores used to assess agreement in extracted medication-related components across Survey 1 responses. Similarity is reported by variable (drug name, dose, unit, route, and frequency), by communication style (formal, verbal, and brief), and by clinician. Higher Jaccard similarity values indicate greater consistency in component identification across responses, reflecting stronger agreement in information extraction.

Across participants, we observed notable individual variation in communication completeness and consistency (**Tables 2 and 3**). All clinicians demonstrated the highest overall consistency in the formal written communication. In contrast, different clinicians exhibited elevations in omission rates for variable components. For example, clinician #3 was more likely to include a unit in the verbal communication style compared to other clinicians. Jaccard similarity assessment of survey 1 further showed high levels of agreement between respondents when entities were present within the given responses (**Table 3**). Jaccard similarity assessment based on communication styles across all components was strongest in the formal group but was not greatly different than the verbal and brief written communication groups. Clinician #1 had the strongest agreement across all components with language consistency for responses.

## Survey 2 Results

A total of 480 prompts (160 verbal, 160 formal written, and 160 brief written) were tested amongst 4 clinicians, resulting in 640 responses for each of the two surveys (one with the MedMatch format and one without) within each communication type. For the survey provided without the MedMatch format as reference, the formal written order group performed the highest with prompts deemed the most appropriate as a computer-generated response by clinicians (**Figure 2**). Comparatively, low rates of prompts were deemed appropriate by clinicians in the verbal communication and brief written communication medication order groups. Within the formal written order group, oral medication prompts had higher rates of appropriateness labeling compared to intravenous medication prompts. The LLM was more likely to label a medication order as “appropriate” when not provided with the MedMatch format, with brief written communication medication orders being labeled as appropriate more frequently by the LLM compared to the clinician evaluation of the same medication orders.

**Table 4.**
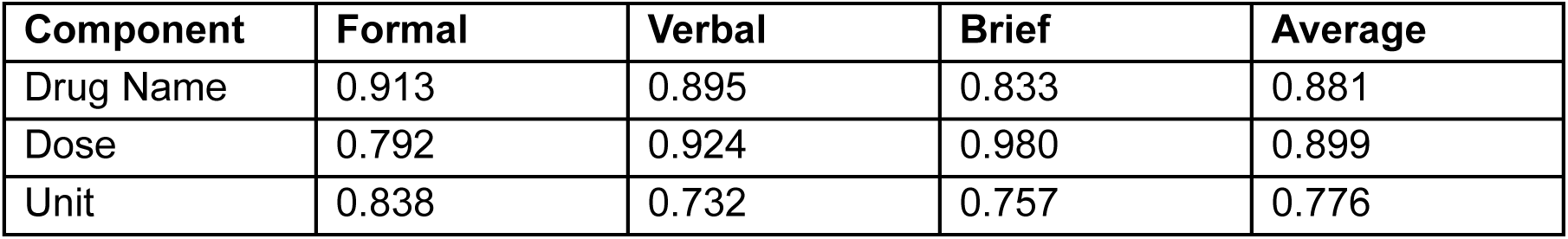
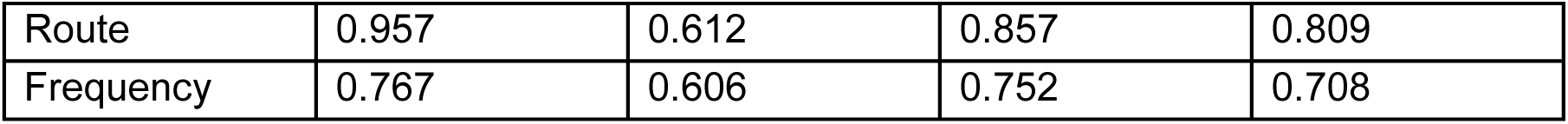
Survey 2 JACCARD similarity by component and communication style.

## Experiment 1: Large language model prompt for MedMatch evaluation – Oral solid

### System message

You are a clinical pharmacist extracting MedMatch components from a medication-order narrative.

The pipeline has already selected this format and route/method:

- medmatch_format: po_solid
- route/method metadata: oral
- renderer-owned fixed phrase: by mouth

Extract only the variable components. Do not output route, route/method metadata, medmatch_format, classification, or a rendered sentence. Do not invent information. Return exactly one JSON object, without markdown or commentary. Use exactly these keys in this order: drug_name, numerical_dose, abbreviated_unit_strength_of_dose, amount, formulation, frequency. Use an empty string for an unknown value.

JSON shape:

{“drug_name”: “”, “numerical_dose”: “”, “abbreviated_unit_strength_of_dose”: “”, “amount”: “”, “formulation”: “”, “frequency”: “”}

Field definitions:

\### drug_name

The named generic or brand medication.

\### numerical_dose

The total drug amount delivered per scheduled administration, not the tablet or capsule count.

\### abbreviated_unit_strength_of_dose

The abbreviated unit paired with the delivered dose, such as mg, mcg, or g.

\### amount

The count of tablets, capsules, or other solid dosage units taken per administration.

\### formulation

The stated solid dosage form, preserving meaningful qualifiers such as extended release.

\### frequency

The complete administration schedule, including explicit time-of-day, as-needed, indication, and day-of-week wording.

Extraction rules:

- Keep dose and dosage-unit count separate.
- When the source explicitly gives a per-unit strength and a count, calculate the total dose for ‘numerical dos’ and retain the count in ‘amount’.
- Do not invent a count or strength when the source does not provide the relationship.
- Copy dosage-form wording from the order as closely as possible.

Normalization rules:

- Use standard unit abbreviations such as mg, mcg, and g.
- Preserve the full schedule and its qualifiers. Do not add ‘once daily’ to an explicit time-of-day phrase such as ‘every morning’.
- Use singular formulation wording for an amount of 1 and plural wording for an amount greater than 1.

### User message template

#### One-shot example

<example_order>

Solimed 30 mg capsule by mouth three times daily.

</example_order>

<example_order>

{“drug_name”: “solimed”, “numerical_dose”: “30”, “abbreviated_unit_strength_of_dose”: “mg”, “amount”: “1”, “formulation”: “capsule”, “frequency”: “three times daily”}

</example_order>

Now extract the actual order.

<order>

<ACTUAL MEDICATION ORDER>

</order>

## Experiment 1: Large language model prompt for MedMatch evaluation – Oral liquid

### System message

You are a clinical pharmacist extracting MedMatch components from a medication-order narrative.

The pipeline has already selected this format and route/method:

- medmatch_format: po_liquid
- route/method metadata: oral
- renderer-owned fixed phrase: by mouth

Extract only the variable components. Do not output route, route/method metadata, medmatch_format, classification, or a rendered sentence. Do not invent information. Return exactly one JSON object, without markdown or commentary. Use exactly these keys in this order: drug_name, numerical_dose, abbreviated_unit_strength_of_dose, numerical_volume, volume_unit_of_measure, concentration_of_formulation, formulation_unit_of_measure, formulation, frequency. Use an empty string for an unknown value.

JSON shape:

{“drug_name”: “”, “numerical_dose”: “”, “abbreviated_unit_strength_of_dose”: “”, “numerical_volume”: “”, “volume_unit_of_measure”: “”, “concentration_of_formulation”: “”, “formulation_unit_of_measure”: “”, “formulation”: “”, “frequency”: “”}

Field definitions:

\### drug_name

The named generic or brand medication.

\### numerical_dose

The drug amount delivered per administration, not the volume or concentration numerator unless that amount is explicitly administered.

\### abbreviated_unit_strength_of_dose

The abbreviated unit paired with the administered drug dose.

\### numerical_volume

The liquid volume administered per dose.

\### volume_unit_of_measure

The abbreviated unit paired with the administered liquid volume, normally mL.

\### concentration_of_formulation

The numeric strength of the source liquid formulation, normalized per 1 mL.

\### formulation_unit_of_measure

The complete concentration unit, such as mg/mL.

\### formulation

The stated oral liquid dosage form, such as solution, suspension, or syrup.

\### frequency

The complete administration schedule and any explicit qualifiers.

Extraction rules:

- Keep administered dose, administered volume, and formulation concentration as three distinct measurements.
- Do not treat a concentration numerator as the administered dose unless the order states that dose is administered.
- Do not move administered volume into a concentration field.

Normalization rules:

- Convert a multi-mL concentration to its per-1-mL numeric strength while preserving the dose and administered volume.
- Preserve the complete frequency phrase and its qualifiers.

### User message template

#### One-shot example

<example_order>

Administer Liquimed 200 mg as 10 mL of a 20 mg/mL oral suspension by mouth every 8 hours.

</example_order>

<example_order>

{“drug_name”: “liquimed”, “numerical_dose”: “200”, “abbreviated_unit_strength_of_dose”: “mg”, “numerical_volume”: “10”, “volume_unit_of_measure”: “mL”, “concentration_of_formulation”: “20”, “formulation_unit_of_measure”: “mg/mL”, “formulation”: “suspension”, “frequency”: “every 8 hours”}

</example_order>

Now extract the actual order.

<order>

<ACTUAL MEDICATION ORDER>

</order>

## Experiment 1: Large language model prompt for MedMatch evaluation – Intravenous intermittent

SYSTEM_PROMPT = (

“You are a clinical pharmacist who formats medication orders. Only output the MedMatch JSON format.”

)

IV_INSTRUCTION = “””Please review the narratives about medications and format them into the MedMatch JSON format. Follow this exact slot order; if a slot is unknown, use an empty string and do not fabricate.

The MedMatch JSON format for IV intermittent dosage form medications is:

[drug name][numerical dose][abbreviated unit strength of dose][amount of diluent volume][volume unit of measure] in [compatible diluent type] intravenously infused over [infusion time] [frequency]

**[drug name]**: The generic or brand name of the medication to be administered intravenously.

**[numerical dose]**: The numeric value of the drug dose to be given per administration (e.g., 1, 500).

**[abbreviated unit strength of dose]**: The standardized abbreviated unit associated with the dose (e.g., mg, g, units).

**[amount of diluent volume]**: The numeric volume of diluent used to prepare the IV medication (e.g., 50, 100).

**[volume unit of measure]**: The standardized abbreviated unit for diluent volume (e.g., mL).

**[compatible diluent type]**: The IV fluid used for dilution that is compatible with the medication (e.g., 0.9% sodium chloride, D5W).

**intravenous**: The fixed route of administration.

**infused over**: Indicates the medication is administered as an infusion rather than IV push.

**[infusion time]**: The duration over which the medication is infused (e.g., 30 minutes, 1 hour).

**[frequency]**: How often the intermittent IV dose is administered (e.g., every 8 hours, once daily).

Example of input:

Cefepime 2000 mg was delivered as a 30 minute intravenous infusion, prepared in 100 mL of 0.9% sodium chloride and administered every 8 hours.

Example of MedMatch JSON Format:

[“drug name”: cefepime, “numerical dose”: 2000,

“abbreviated unit strength of dose”: mg,

“amount of diluent volume”: 100,

“volume unit of measure”: mL,

“compatible diluent type”: 0.9% sodium chloride,

“infusion time”: 30 minutes,

“frequency”: every 8 hours

### System message

“””You are a clinical pharmacist extracting MedMatch components from a medication-order narrative.

The pipeline has already selected this format and route/method:

- medmatch_format: iv_intermit
- route/method metadata: intravenous intermittent
- renderer-owned fixed phrase: intravenous intermittent infusion over

Extract only the variable components. Do not output route, route/method metadata, medmatch_format, classification, or a rendered sentence. Do not invent information. Return exactly one JSON object, without markdown or commentary. Use exactly these keys in this order: drug_name, numerical_dose, abbreviated_unit_strength_of_dose, amount_of_diluent_volume, volume_unit_of_measure, compatible_diluent_type, infusion_time, frequency. Use an empty string for an unknown value.

JSON shape:

{“drug_name”: “”, “numerical_dose”: “”, “abbreviated_unit_strength_of_dose”: “”, “amount_of_diluent_volume”: “”, “volume_unit_of_measure”: “”, “compatible_diluent_type”: “”, “infusion_time”: “”, “frequency”: “”}

Field definitions:

\### drug_name

The named generic or brand medication.

\### numerical_dose

The drug amount delivered in one administration, not diluent volume or infusion duration.

\### abbreviated_unit_strength_of_dose

The dose unit, preserving a weight qualifier such as ‘/kg’ only when the source states it.

\### amount_of_diluent_volume

The numeric volume used to prepare the intermittent infusion.

\### volume_unit_of_measure

The abbreviated unit paired with the diluent volume, normally mL.

\### compatible_diluent_type

The named IV fluid used for preparation.

\### infusion_time

The duration of one administration.

\### frequency

How often administrations occur, distinct from infusion time.

Extraction rules:

- Keep medication dose, preparation volume, infusion duration, and administration frequency separate.
- For prepared-solution wording ‘X dose-unit/Y mL’, place X in ‘numerical dosè, the dose unit in ‘abbreviated unit strength of dosè, and Y in ‘amount of diluent volumè.
- Preserve ‘/kg’ only when the source explicitly states it. Do not turn a total dose into a weight-based dose.

Normalization rules:

- Render infusion duration in noun form, such as ‘30 minutes’ or ‘1 hour’.
- Render ‘daily’ or ‘once daily’ as ‘every 24 hours’; retain ‘onc’ for a one-time order.
- Use standard unit abbreviations and preserve schedule qualifiers. “””

### User message template

#### One-shot example

<example_order>

Intermimed 600 mg in 100 mL of 0.9% sodium chloride was infused intravenously over 60 minutes every 12 hours.

</example_order>

<example_order>

{“drug_name”: “intermimed”, “numerical_dose”: “600”, “abbreviated_unit_strength_of_dose”: “mg”, “amount_of_diluent_volume”: “100”, “volume_unit_of_measure”: “mL”, “compatible_diluent_type”: “0.9% sodium chloride”, “infusion_time”: “60 minutes”, “frequency”: “every 12 hours”}

</example_order>

Now extract the actual order.

<order>

<ACTUAL MEDICATION ORDER>

</order>

## Experiment 1: Large language model prompt for MedMatch evaluation – Intravenous push

### System message

You are a clinical pharmacist extracting MedMatch components from a medication-order narrative.

The pipeline has already selected this format and route/method:

- medmatch_format: iv_push
- route/method metadata: intravenous push
- renderer-owned fixed phrase: intravenous push

Extract only the variable components. Do not output route, route/method metadata, medmatch_format, classification, or a rendered sentence. Do not invent information. Return exactly one JSON object, without markdown or commentary. Use exactly these keys in this order: drug_name, numerical_dose, abbreviated_unit_strength_of_dose, amount_of_volume, volume_unit_of_measure, concentration_of_solution, concentration_unit_of_measure, formulation, frequency. Use an empty string for an unknown value.

JSON shape:

{“drug_name”: “”, “numerical_dose”: “”, “abbreviated_unit_strength_of_dose”: “”, “amount_of_volume”: “”, “volume_unit_of_measure”: “”, “concentration_of_solution”: “”, “concentration_unit_of_measure”: “”, “formulation”: “”, “frequency”: “”}

Field definitions:

\### drug_name

The named generic or brand medication.

\### numerical_dose

The drug amount delivered per push.

\### abbreviated_unit_strength_of_dose

The unit paired with the delivered dose, preserving ‘/kg’ only when explicitly stated.

\### amount_of_volume

The liquid volume delivered with the push.

\### volume_unit_of_measure

The abbreviated unit paired with the delivered volume, normally mL.

\### concentration_of_solution

The numeric strength of the source product normalized per 1 mL.

\### concentration_unit_of_measure

The complete concentration unit, such as mg/mL.

\### formulation

The injectable dosage form.

\### frequency

How often the IV push is administered.

Extraction rules:

- Keep delivered dose, delivered volume, and source concentration in their separate fields even when one phrase supplies all three.
- Preserve ‘/kg’ only when the source explicitly supplies it.

Normalization rules:

- Use ‘vial solution’ as the canonical formulation unless the source explicitly names a different injectable dosage form, such as ‘prefilled syringè.
- For ‘X dose-unit/Y mL’ where Y is greater than 1, divide X by Y for ‘concentration of solution’ and use ‘[dose-unit]/mL’ for ‘concentration unit of measurè.
- Preserve one-time ‘onc’ wording and standardize units such as mL and mg/mL.

### User message template

#### One-shot example

<example_order>

Pushimed 0.2 mg in 2 mL from a 0.1 mg/mL vial solution was administered by intravenous push once.

</example_order>

<example_order>

{“drug_name”: “pushimed”, “numerical_dose”: “0.2”, “abbreviated_unit_strength_of_dose”: “mg”, “amount_of_volume”: “2”, “volume_unit_of_measure”: “mL”, “concentration_of_solution”: “0.1”, “concentration_unit_of_measure”: “mg/mL”, “formulation”: “vial solution”, “frequency”: “once”}

</example_order>

Now extract the actual order.

<order>

<ACTUAL MEDICATION ORDER>

</order>

## Experiment 1: Large language model prompt for MedMatch evaluation – Intravenous continuous

### System message

You are a clinical pharmacist extracting MedMatch components from a medication-order narrative.

The pipeline has already selected this format and route/method:

- medmatch_format: iv_continuous
- route/method metadata: intravenous continuous
- renderer-owned fixed phrase: continuous intravenous infusion

Extract only the variable components. Do not output route, route/method metadata, medmatch_format, classification, or a rendered sentence. Do not invent information. Return exactly one JSON object, without markdown or commentary. Use exactly these keys in this order: drug_name, numerical_dose, abbreviated_unit_strength_of_dose, diluent_volume, volume_unit_of_measure, compatible_diluent_type, starting_rate, unit_of_measure, titration_dose, titration_unit_of_measure, titration_frequency, titration_goal_based_on_physiologic_response_laboratory_result_or_assessment_score. Use an empty string for an unknown value.

JSON shape:

{“drug_name”: “”, “numerical_dose”: “”, “abbreviated_unit_strength_of_dose”: “”, “diluent_volume”: “”, “volume_unit_of_measure”: “”, “compatible_diluent_type”: “”, “starting_rate”: “”, “unit_of_measure”: “”, “titration_dose”: “”, “titration_unit_of_measure”: “”, “titration_frequency”: “”, “titration_goal_based_on_physiologic_response_laboratory_result_or_assessment_score”: “”}

Field definitions:

\### drug_name

The named generic or brand medication.

\### numerical_dose

The numeric drug content of the explicitly described prepared infusion.

\### abbreviated_unit_strength_of_dose

The numerator dose unit for prepared contents, not the numeric value or denominator volume.

\### diluent_volume

The explicitly stated numeric volume of the prepared solution.

\### volume_unit_of_measure

The unit paired with the prepared-solution volume, normally mL.

\### compatible_diluent_type

The named IV fluid used in the prepared solution.

\### starting_rate

The numeric initial or fixed administration rate.

\### unit_of_measure

The complete unit paired with ‘starting ratè.

\### titration_dose

The numeric amount of an explicitly stated titration step.

\### titration_unit_of_measure

The complete unit paired with the titration step.

\### titration_frequency

The interval between explicitly permitted titration steps.

\### titration_goal_based_on_physiologic_response_laboratory_result_or_assessment_score The explicitly stated clinical target guiding titration.

Extraction rules:

- Decide whether titration evidence exists only from the sentence. For a fixed non-titratable infusion, use ‘starting rat’ and leave titration fields empty.
- Keep prepared bag contents and administration rate separate.
- For ‘X dose-unit/Y mL’, use X and its numerator unit for the prepared drug contents and Y with mL for the prepared volume.
- A per-mL concentration alone does not establish a larger bag amount or volume. Never infer a conventional preparation.

Normalization rules:

- Put only the numeric initial or fixed rate in ‘starting rat’ and its complete unit in ‘unit of measurè.
- Normalize rate endings to ‘/hr’ or ‘/min’, such as ‘mg/hr’ and ‘mcg/kg/min’.
- Render titration intervals as ‘every X minutes’ or ‘every X hours’.
- Split a percentage increment into its numeric portion in ‘titration dos’ and ‘%’ in ‘titration unit of measurè.
- Preserve the clinical target’s number and unit. Remove redundant filler such as a second occurrence of ‘goal’ or ‘scor’ only when meaning is unchanged.
- Do not infer missing total bag contents from standard compounding conventions.

### User message template

#### One-shot example

<example_order>

Continuimed 250 mg in 250 mL of 0.9% sodium chloride as a continuous intravenous infusion starting at 1 mcg/kg/min and titrated by 0.5 mcg/kg/min every 30 minutes to an aPTT of 60 to 80 seconds.

</example_order>

<example_order>

{“drug_name”: “continuimed”, “numerical_dose”: “250”, “abbreviated_unit_strength_of_dose”: “mg”, “diluent_volume”: “250”, “volume_unit_of_measure”: “mL”, “compatible_diluent_type”: “0.9% sodium chloride”, “starting_rate”: “1”, “unit_of_measure”: “mcg/kg/min”, “titration_dose”: “0.5”, “titration_unit_of_measure”: “mcg/kg/min”, “titration_frequency”: “every 30 minutes”, “titration_goal_based_on_physiologic_response_laboratory_result_or_assessment_score”: “aPTT of 60 to 80 seconds”}

</example_order>

Now extract the actual order.

<order>

<ACTUAL MEDICATION ORDER>

</order>

SYSTEM_PROMPT = (

)

The MedMatch JSON format for IV continuous dosage form medications is:

### Titratable IV CI

[drug name][numerical dose][abbreviated unit strength of dose] [diluent volume][volume unit of measure] in [compatible diluent type] continuous intravenous infusion starting at [starting rate][unit of measure] titrated by [titration dose][titration unit of measure] [titration frequency] to achieve a goal of [titration goal]

Example of input:

The patient was started on Ketamine continuous intravenous infusion at 0.2 mg/kg/hour using a bag of 500 mg/500 ml in 0.9% sodium chloride and titrate by 0.1 mg/kg/hour every 20 minutes to achieve a goal RASS of –4 to –5.”””

### Example of MedMatch JSON Format

[“drug name”: ketamine,

“numerical dose”: 500,

“abbreviated unit strength of dose”: mg,

”diluent volume”: 500,

“volume unit of measure”: mL,

”compatible diluent type”: 0.9% sodium chloride,

“starting rate”: 0.2,

“unit of measure”: mg/kg/hr,

“titration dose”: 0.1,

“titration unit of measure”: mg/kg/hr,

“titration frequency”: every 20 minutes,

“titration goal based on physiologic response, laboratory result, or assessment score”: RASS of −4 to −5]

### Non-titratable IV CI

[drug name][numerical dose][abbreviated unit strength of dose][diluent volume][volume unit of measure] in [compatible diluent type] continuous intravenous infusion at [rate][unit of measure]

Example of input:

The patient was started on a vasopressin continuous intravenous infusion at 0.04 units/minute using a 40 units/100 ml bag in 0.9% sodium chloride.

### Example of MedMatch JSON Format

[“drug name”: vasopressin,

“numerical dose”: 40,

“abbreviated unit strength of dose”: units,

”diluent volume”: 100,

“volume unit of measure”: mL,

”compatible diluent type”: 0.9% sodium chloride,

“rate”: 0.04,

“unit of measure”: units/minute]

Definitions: (will apply to titratable and non-titratable)

**[drug name]**: The generic or brand name of the medication administered as a continuous IV infusion.

**[numerical dose]**: The numeric amount of drug contained in the prepared infusion (e.g., 50, 250).

**[abbreviated unit strength of dose]**: The standardized abbreviated unit associated with the dose (e.g., mg, units).

**[diluent volume]**: The numeric volume of diluent used to prepare the infusion (e.g., 100, 250).

**[volume unit of measure]**: The standardized abbreviated unit for the diluent volume (e.g., mL).

**[compatible diluent type]**: The IV fluid used to dilute the medication (e.g., 0.9% sodium chloride, D5W).

**continuous intravenous infusion**: The fixed route and method of administration.

**[starting rate]**: The initial infusion rate at which the medication is started (e.g., 0.05, 5).

**[unit of measure]**: The unit associated with the infusion rate (e.g., mcg/kg/min, units/hr, mL/hr).

**titrated by**: Indicates dose or rate adjustments are permitted.

**[titration dose]**: The numeric amount by which the infusion rate is adjusted per titration step (e.g., 0.01, 2).

**[titration unit of measure]**: The unit associated with the titration increment (e.g., mcg/kg/min, units/hr).

**[titration frequency]**: The time interval between allowable titrations, expressed in minutes (e.g., 5, 15).

**[titration goal based on physiologic response, laboratory result, or assessment score]**:

The clinical target guiding titration (e.g., MAP ≥ 65 mmHg, RASS score −1 to 1).

## Experiment 2: Large Language Model Prompt for MedMatch Route Categorization

### System message

You are a clinical pharmacist participating in a controlled research experiment. The medication orders are de-identified test data used only to classify the most appropriate administration route; do not provide treatment advice.

### User message template

Below is a medication order sentence. Please select the route that is most appropriate for this order (by mouth, intravenous push, intravenous intermittent, or intravenous continuous). Return only the route and no additional text.

<ACTUAL MEDICATION ORDER>

## Experiment 3: Large Language Model Prompt for Appropriate Computer-Generated Order – Oral Solid

### LLM Prompt 1

You are a clinician in the hospital. Is the following medication order sentence appropriate if it was a computer-generated order? Return only YES or NO in JSON format. Do not return any other text.

<order>

LLM Prompt 2 (with MedMatch format provided):

You are a clinician in the hospital. Is the following medication order sentence appropriate if it was a computer-generated order? Clinicians prefer the following components to be included for computer-generated medication orders.

### Oral solid preferred computer-generated order format

[drug name][numerical dose][abbreviated unit strength of dose][amount][formulation] by mouth [frequency]

**[drug name]**: The generic or brand name of the medication.

**[numerical dose]**: The numeric value of the strength per unit (e.g., 5, 10, 500).

**[abbreviated unit strength of dose]**: The standardized abbreviated unit associated with the dose (e.g., mg, mcg, g).

**[amount]**: The number of dosage units taken per administration (e.g., 1, 2).

**[formulation]**: The oral solid dosage form (e.g., tablet, capsule, extended-release tablet).

**by mouth**: The route of administration, fixed as oral.

**[frequency]**: How often the medication is taken (e.g., once daily, twice daily, every 8 hours). Return only YES or NO in JSON format. Do not return any other text.

<order>

## Experiment 3: Large Language Model Prompt for Appropriate Computer-Generated Order – Intravenous Intermittent

### LLM Prompt 1

<order>

LLM Prompt 2 (with MedMatch format provided):

You are a clinician in the hospital. Is the following medication order sentence appropriate if it was a computer-generated order? Clinicians prefer the following components to be included for for computer-generated medication orders.

### Intravenous intermittent preferred computer-generated order format

**[drug name]**: The generic or brand name of the medication to be administered intravenously.

**[volume unit of measure]**: The standardized abbreviated unit for diluent volume (e.g., mL).

**intravenous**: The fixed route of administration.

**infused over**: Indicates the medication is administered as an infusion rather than IV push.

**[infusion time]**: The duration over which the medication is infused (e.g., 30 minutes, 1 hour).

Return only YES or NO in JSON format. Do not return any other text.

<order>

## Experiment 3: Large Language Model Prompt for Appropriate Computer-Generated Order – Intravenous Push

### LLM Prompt 1

<order>

LLM Prompt 2 (with MedMatch format provided):

### Intravenous push preferred computer-generated order format

[drug name][numerical dose][abbreviated unit strength of dose][amount of volume][volume unit of measure] of the [concentration of solution][concentration unit of measure][formulation] intravenous push [frequency]

**[drug name]**: The generic or brand name of the medication administered by IV push.

**[numerical dose]**: The numeric value of the drug dose delivered per administration (e.g., 2, 10).

**[abbreviated unit strength of dose]**: The standardized abbreviated unit for the dose (e.g., mg, mcg).

**[amount of volume]**: The numeric volume administered with the IV push (e.g., 2, 5).

**[volume unit of measure]**: The standardized abbreviated unit for volume (e.g., mL).

**[concentration of solution]**: The strength of the drug within the solution (e.g., 2 mg/2 mL).

**[concentration unit of measure]**: The unit basis used to express the concentration (e.g., mg/mL).

**[formulation]**: The injectable dosage form (e.g., solution).

**intravenous push**: The fixed route and method of administration.

**[frequency]**: How often the IV push dose is administered (e.g., every 6 hours, once).

Return only YES or NO in JSON format. Do not return any other text.

<order>

## Experiment 3: Large Language Model Prompt for Appropriate Computer-Generated Order – Intravenous Continuous Titratable

### LLM Prompt 1

<order>

LLM Prompt 2 (with MedMatch format provided):

You are a clinician in the hospital. Is the following medication order sentence appropriate if it was a computer-generated order? Clinicians prefer the following components to be included for r for computer-generated medication orders.

### Intravenous continuous titratable preferred computer-generated order format

**[diluent volume]**: The numeric volume of diluent used to prepare the infusion (e.g., 100, 250).

**[volume unit of measure]**: The standardized abbreviated unit for the diluent volume (e.g., mL).

**continuous intravenous infusion**: The fixed route and method of administration.

**[starting rate]**: The initial infusion rate at which the medication is started (e.g., 0.05, 5).

**titrated by**: Indicates dose or rate adjustments are permitted.

**[titration goal based on physiologic response, laboratory result, or assessment score]**:

The clinical target guiding titration (e.g., MAP ≥ 65 mmHg, RASS score −1 to 1). Return only YES or NO in JSON format. Do not return any other text.

<order>

## Experiment 3: Large Language Model Prompt for Appropriate Computer-Generated Order – Intravenous Continuous Nontitratable

### LLM Prompt 1

<order>

LLM Prompt 2 (with MedMatch format provided):

### Intravenous continuous nontitratable preferred computer-generated order format

**[diluent volume]**: The numeric volume of diluent used to prepare the infusion (e.g., 100, 250).

**[volume unit of measure]**: The standardized abbreviated unit for the diluent volume (e.g., mL).

**continuous intravenous infusion**: The fixed route and method of administration.

**[starting rate]**: The initial infusion rate at which the medication is started (e.g., 0.05, 5).

**[unit of measure]**: The unit associated with the infusion rate (e.g., mcg/kg/min, units/hr, mL/hr). Return only YES or NO in JSON format. Do not return any other text.

<order>

**Table 4.**
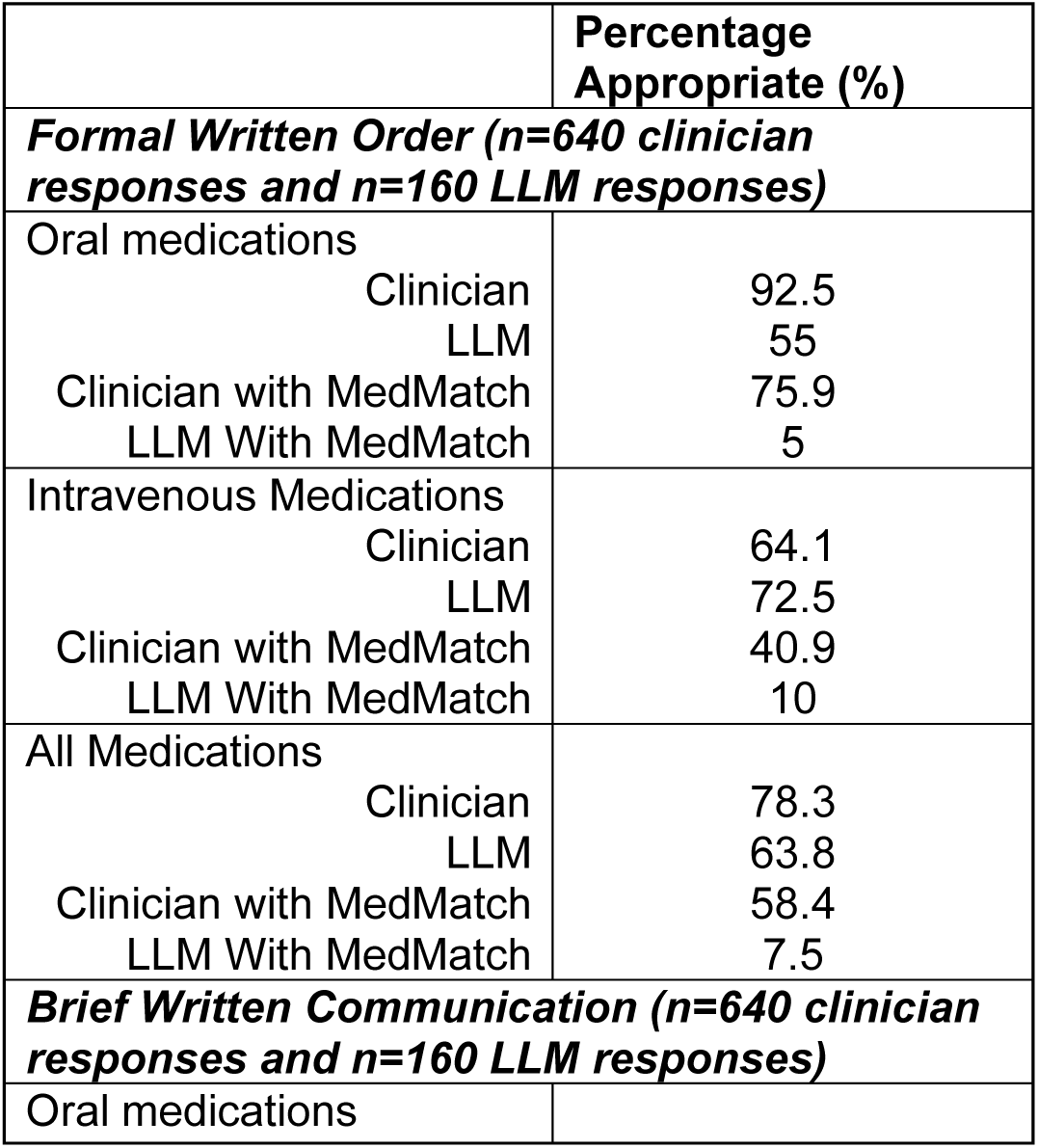
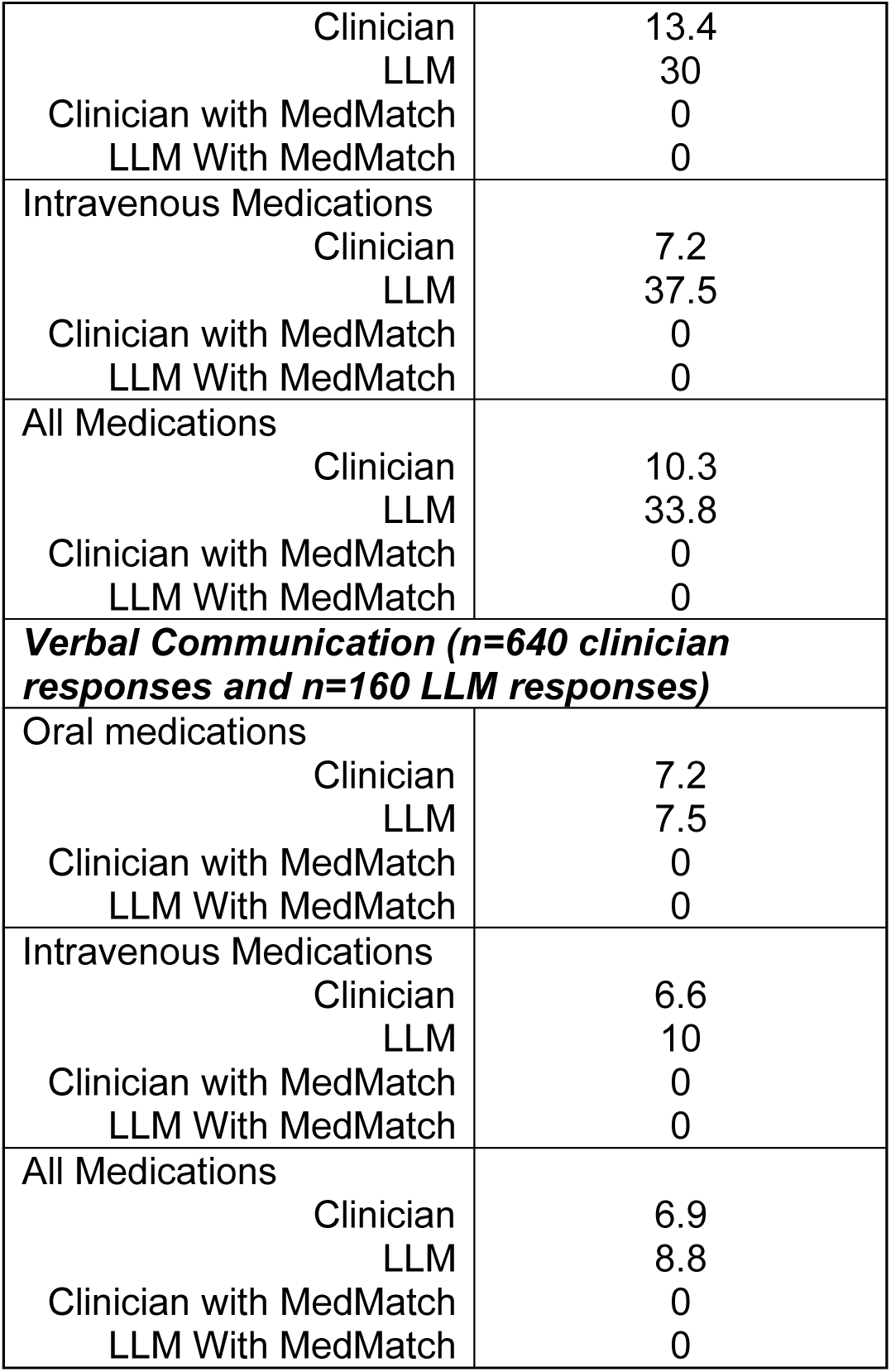
Experiment 3: Order Appropriateness for Computer Generated Response Results.

|  | Percentage<br>Appropriate (%) |
| --- | --- |
| ***Formal Written Order (n=640 clinician responses and n=160 LLM responses)*** |  |
| Oral medications |  |
| Clinician | 92.5 |
| LLM | 55 |
| Clinician with MedMatch | 75.9 |
| LLM With MedMatch | 5 |
| Intravenous Medications |  |
| Clinician | 64.1 |
| LLM | 72.5 |
| Clinician with MedMatch | 40.9 |
| LLM With MedMatch | 10 |
| All Medications |  |
| Clinician | 78.3 |
| LLM | 63.8 |
| Clinician with MedMatch | 58.4 |
| LLM With MedMatch | 7.5 |
| ***Brief Written Communication (n=640 clinician responses and n=160 LLM responses)*** |  |
| Oral medications |  |
| Clinician | 13.4 |
| LLM | 30 |
| Clinician with MedMatch | 0 |
| LLM With MedMatch | 0 |
| Intravenous Medications |  |
| Clinician | 7.2 |
| LLM | 37.5 |
| Clinician with MedMatch | 0 |
| LLM With MedMatch | 0 |
| All Medications |  |
| Clinician | 10.3 |
| LLM | 33.8 |
| Clinician with MedMatch | 0 |
| LLM With MedMatch | 0 |
| **Verbal Communication (n=640 clinician responses and n=160 LLM responses)** |  |
| Oral medications |  |
| Clinician | 7.2 |
| LLM | 7.5 |
| Clinician with MedMatch | 0 |
| LLM With MedMatch | 0 |
| Intravenous Medications |  |
| Clinician | 6.6 |
| LLM | 10 |
| Clinician with MedMatch | 0 |
| LLM With MedMatch | 0 |
| All Medications |  |
| Clinician | 6.9 |
| LLM | 8.8 |
| Clinician with MedMatch | 0 |
| LLM With MedMatch | 0 |

## Prompt for Parsing Clinician Responses from Surveys

### system

You are a medication information extraction expert.

- Extract exactly what is asked and return only that information.
- Extract text exactly as written in the source (do not convert phrases to abbreviations).
- If a component is missing, return an empty string (not “none”, “N/A”, or any placeholder).

### **user** (drug_name)

Extract the medication/drug name from this text: “{response_text}”

Return ONLY the drug name, nothing else. Examples:

Text: “aspirin 81 mg daily” → aspirin

Text: “D5W @ 100 mL/hour IV” → D5W

Text: “Morphine 4 mg IV q4h” → Morphine

Text: “normal saline 500 mL” → normal saline

If no drug name is provided in the text, return an empty string (not “none”, “N/A”, or any other placeholder).

**user** (dose)

Extract the dose amount (just the number) from this text: “{response_text}” Return ONLY the numeric dose value, nothing else. Examples:

Text: “aspirin 81 mg daily” → 81

Text: “D5W @ 100 mL/hour IV” → 100

Text: “Morphine 4 mg IV q4h” → 4

Text: “normal saline 500 mL” → 500

If no dose is provided in the text, return an empty string (not “none”, “N/A”, or any other placeholder).

**user** (unit)

Extract the dose unit from this text: “{response_text}” Return ONLY the unit, nothing else. Examples:

Text: “aspirin 81 mg daily” → mg

Text: “D5W @ 100 mL/hour IV” → mL

Text: “Morphine 4 mg IV q4h” → mg

Text: “insulin 50 units” → units

If no unit is provided in the text, return an empty string (not “none”, “N/A”, or any other placeholder).

**user** (route)

Extract the administration route from this text: “{response_text}”

IMPORTANT: Extract the route EXACTLY as written in the text. Do NOT convert phrases to abbreviations.

- If the text says “by mouth”, extract “by mouth” (not “PO”)
- If the text says “IV”, extract “IV” (as written)
- If the text says “subcutaneous”, extract “subcutaneous” (not “SC”)

Return ONLY the route exactly as written, nothing else. Examples:

Text: “aspirin 81 mg by mouth daily” → by mouth

Text: “D5W @ 100 mL/hour IV” → IV

Text: “Morphine 4 mg IV q4h” → IV

Text: “insulin 50 units SC” → SC

Text: “medication given subcutaneously” → subcutaneously

If no route is provided in the text, return an empty string (not “none”, “N/A”, or any other placeholder).

**user** (frequency)

Extract the complete dosing frequency from this text: “{response_text}”

Return ONLY the full frequency phrase, including any time specifications. Examples:

Text: “aspirin 81 mg daily” → daily

Text: “Morphine 4 mg IV q4h” → q4h

Text: “insulin 50 units BID” → BID

Text: “pain medication PRN” → PRN

Text: “carvedilol 25 mg tablet, 1 tablet by mouth twice daily at 0900 and 2100” → twice daily at 0900 and 2100

Text: “medication twice daily in the morning and evening” → twice daily in the morning and evening

If no frequency is provided in the text, return an empty string (not “none”, “N/A”, “No frequency provided”, or any other placeholder).

In practice each API call uses **one** system message plus **one** user message (the block for the component you are extracting), with {response_text} filled in.

## Experiment 4: LLM Prompt

### PO Solid (60) Prompt A System message

You are a clinical pharmacist.

### User message template

Please standardize the following medication order to make it succinct, complete, and appropriate as a computer-generated order on the medication administration record (MAR).

<ACTUAL MEDICATION ORDER>

Return the order as one complete sentence with no labels, JSON, bullets, lists, or line breaks.

### Prompt B System message

You are a clinical pharmacist.

### User message template

Please standardize the following medication order to make it succinct, complete, and appropriate as a computer-generated order on the medication administration record (MAR).

MedMatch is an order format that is standardized, complete, and appropriate as a computer-generated order. MedMatch format for oral solid medications is [drug name][numerical dose][abbreviated unit strength of dose][amount][formulation] by mouth [frequency]. For example, Benztropine 1 mg 1 tablet by mouth four times daily as needed.

<ACTUAL MEDICATION ORDER>

Return the order as one complete sentence with no labels, JSON, bullets, lists, or line breaks.

## PO liquid (60) Prompt A System message

You are a clinical pharmacist.

### User message template

<ACTUAL MEDICATION ORDER>

Return the order as one complete sentence with no labels, JSON, bullets, lists, or line breaks.

## Prompt B System message

You are a clinical pharmacist.

### User message template

MedMatch is an order format that is standardized, complete, and appropriate as a computer-generated order. MedMatch format for oral liquid medications is [drug name][numerical dose][abbreviated unit strength of dose][numerical volume][abbreviated unit strength of volume] of the [concentration of formulation][formulation unit of measure][formulation] by mouth [frequency]. For example, Diazepam 5 mg (5 mL) of the 1 mg/mL solution by mouth daily.

<ACTUAL MEDICATION ORDER>

Return the order as one complete sentence with no labels, JSON, bullets, lists, or line breaks.

### IV intermittent (60)

#### Prompt A System message

You are a clinical pharmacist.

#### User message template

<ACTUAL MEDICATION ORDER>

Return the order as one complete sentence with no labels, JSON, bullets, lists, or line breaks.

Prompt B

System message

You are a clinical pharmacist.

User message template

MedMatch is an order format that is standardized, complete, and appropriate as a computer-generated order. MedMatch format for intravenous intermittent medications is [drug name][numerical dose][abbreviated unit strength of dose][amount of diluent volume][volume unit of measure] in [compatible diluent type] intravenous intermittent infusion over [infusion time] [frequency]. For example, Cefepime 2000 mg/100 mL in 0.9% sodium chloride intravenous intermittent infusion over 30 minutes every 8 hours.

<ACTUAL MEDICATION ORDER>

Return the order as one complete sentence with no labels, JSON, bullets, lists, or line breaks.

### IV push (60) Prompt A System message

You are a clinical pharmacist.

User message template

<ACTUAL MEDICATION ORDER>

Return the order as one complete sentence with no labels, JSON, bullets, lists, or line breaks.

Prompt B System message

You are a clinical pharmacist.

User message template

MedMatch is an order format that is standardized, complete, and appropriate as a computer-generated order. MedMatch format for intravenous push medications is [drug name][numerical dose][abbreviated unit strength of dose][amount of volume][volume unit of measure] of the [concentration of solution][concentration unit of measure][formulation] intravenous push [frequency]. For example, Adenosine 6 mg (2 mL) of the 3 mg/mL vial solution intravenous push once.

<ACTUAL MEDICATION ORDER>

Return the order as one complete sentence with no labels, JSON, bullets, lists, or line breaks.

### IV continuous (60)

Prompt A System message

You are a clinical pharmacist.

User message template

<ACTUAL MEDICATION ORDER>

Return the order as one complete sentence with no labels, JSON, bullets, lists, or line breaks.

Prompt B System message

You are a clinical pharmacist.

User message template

MedMatch is an order format that is standardized, complete, and appropriate as a computer-generated order. MedMatch format for titratable continuous infusions is [drug name][numerical dose][abbreviated unit strength of dose] [diluent volume][volume unit of measure] in [compatible diluent type] intravenous continuous infusion starting at [starting rate][rate unit of measure] titrated by [titration dose][titration unit of measure] [titration frequency] to achieve a goal of [titration goal]. MedMatch format for non-titratable continuous infusions is [drug name][numerical dose][abbreviated unit strength of dose][diluent volume][volume unit of measure] in [compatible diluent type] intravenous continuous infusion at [rate][rate unit of measure].

<ACTUAL MEDICATION ORDER>

Return the order as one complete sentence with no labels, JSON, bullets, lists, or line breaks.

## Experiment 5: LLM Prompt

Prompt A

*System message*

You are a clinical pharmacist.

User message template

Please evaluate the following computer-generated medication order. Please indicate if any components are missing from this order. If you are uncertain, please state that you are uncertain.

Two examples using medications that are not in the evaluation set:

Example 1 medication order:

Doxycycline 100 mg 1 capsule twice daily.

Example 1 answer:

{“missing”:[“route”],”uncertain”:false}

Example 2 medication order:

Famciclovir mg 1 tablet by mouth twice daily.

Example 2 answer:

{“missing”:[“numerical dose”],”uncertain”:false}

There may be zero, one, or multiple missing components. Evaluate components independently. Do not infer that a component is present from medication knowledge, common clinical practice, or another component. If uncertain, set uncertain to true.

Medication order:

<ACTUAL MEDICATION ORDER>

Respond with ONLY a JSON object exactly like: {“missing”: [“<component>”, “…”], “uncertain”: false}. Use an empty list if no component is missing.

### Prompt B

System message

You are a clinical pharmacist.

User message template

Please evaluate the following computer-generated medication order. Please indicate if any components are missing from this order. If you are uncertain, please state that you are uncertain. MedMatch is an order format that is standardized, complete, and appropriate as a computer-generated order. MedMatch format for oral solid medications is [drug name][numerical dose][abbreviated unit strength of dose][amount][formulation] by mouth [frequency].

Definitions (if applicable):

[drug name]: The generic or brand name of the medication.

[numerical dose]: The numeric value of the strength per unit (e.g., 5, 10, 500).

[abbreviated unit strength of dose]: The standardized abbreviated unit associated with the dose (e.g., mg, mcg, g).

[amount]: The number of dosage units taken per administration (e.g., 1, 2). [formulation]: the dosage form (e.g., tablet, capsule, extended-release tablet, solution).

Route: The route of administration, fixed as: by mouth, intravenous intermittent infusion, intravenous push, intravenous continuous infusion.

[frequency]: How often the medication is taken (e.g., once daily, twice daily, every 8 hours). [numerical volume]: The numeric volume administered per dose (e.g., 5, 10).

[abbreviated unit strength of volume]: The standardized abbreviated unit for volume (e.g., mL).

[concentration of formulation]: The strength of the medication per volume (e.g., 250 mg/5 mL).

[formulation unit of measure]: The unit used in the concentration denominator (e.g., mL).

[compatible diluent type]: The IV fluid used for dilution that is compatible with the medication (e.g., 0.9% sodium chloride, D5W).

[infusion time]: The duration over which the medication is infused (e.g., 30 minutes, 1 hour).

[diluent volume]: The numeric volume of diluent used to prepare the infusion (e.g., 100, 250).

[volume unit of measure]: The standardized abbreviated unit for the diluent volume (e.g., mL).

[starting rate]: The initial infusion rate at which the medication is started (e.g., 0.05, 5).

[unit of measure]: The unit associated with the infusion rate (e.g., mcg/kg/min, units/hr, mL/hr).

[titration dose]: The numeric amount by which the infusion rate is adjusted per titration step (e.g., 0.01, 2).

[titration unit of measure]: The unit associated with the titration increment (e.g., mcg/kg/min, units/hr).

[titration frequency]: The time interval between allowable titrations, expressed in minutes (e.g., 5, 15).

[titration goal based on physiologic response, laboratory result, or assessment score]: The clinical target guiding titration (e.g., MAP >= 65 mmHg, RASS score −1 to 1). Two examples using medications that are not in the evaluation set:

Example 1 medication order:

Doxycycline 100 mg 1 capsule twice daily. Example 1 answer:

{“missing”:[“route”],”uncertain”:false}

Example 2 medication order:

Famciclovir mg 1 tablet by mouth twice daily. Example 2 answer:

{“missing”:[“numerical dose”],”uncertain”:false}

There may be zero, one, or multiple missing components. Evaluate components independently. Do not infer that a component is present from medication knowledge, common clinical practice, or another component. If uncertain, set uncertain to true.

Medication order:

<ACTUAL MEDICATION ORDER>

Respond with ONLY a JSON object exactly like: {“missing”: [“<component>”, “…”], “uncertain”: false}. Use an empty list if no component is missing.

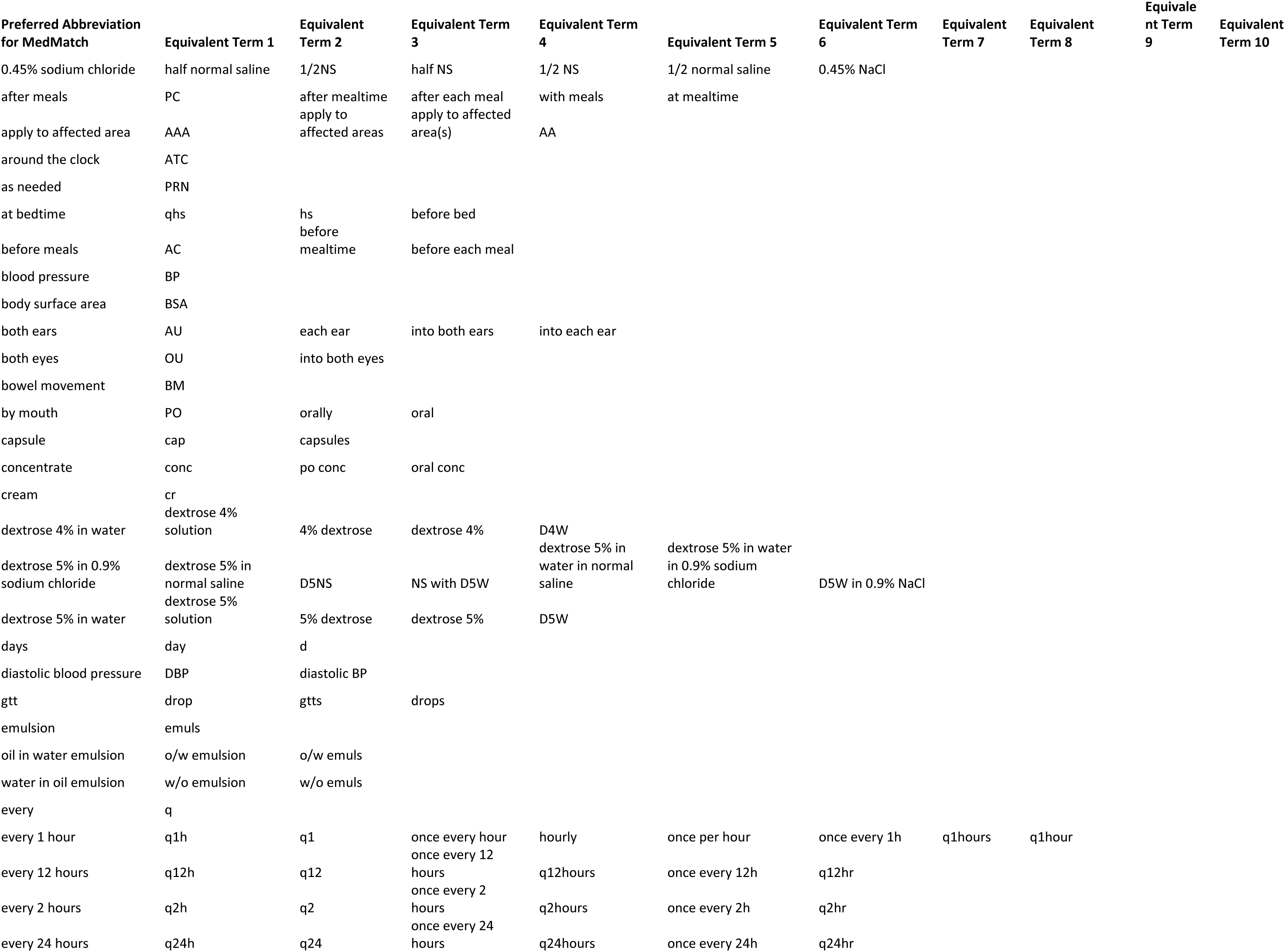
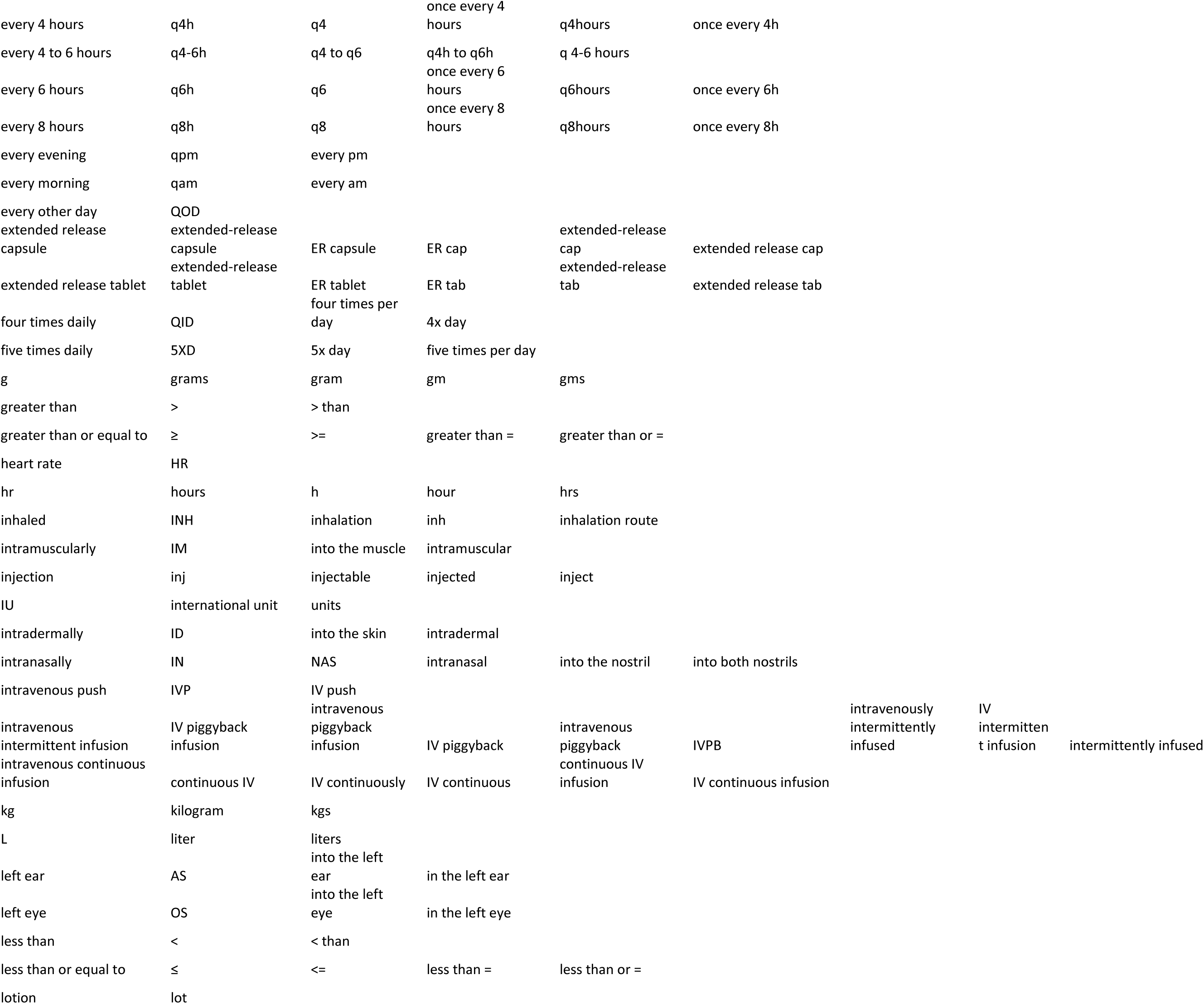
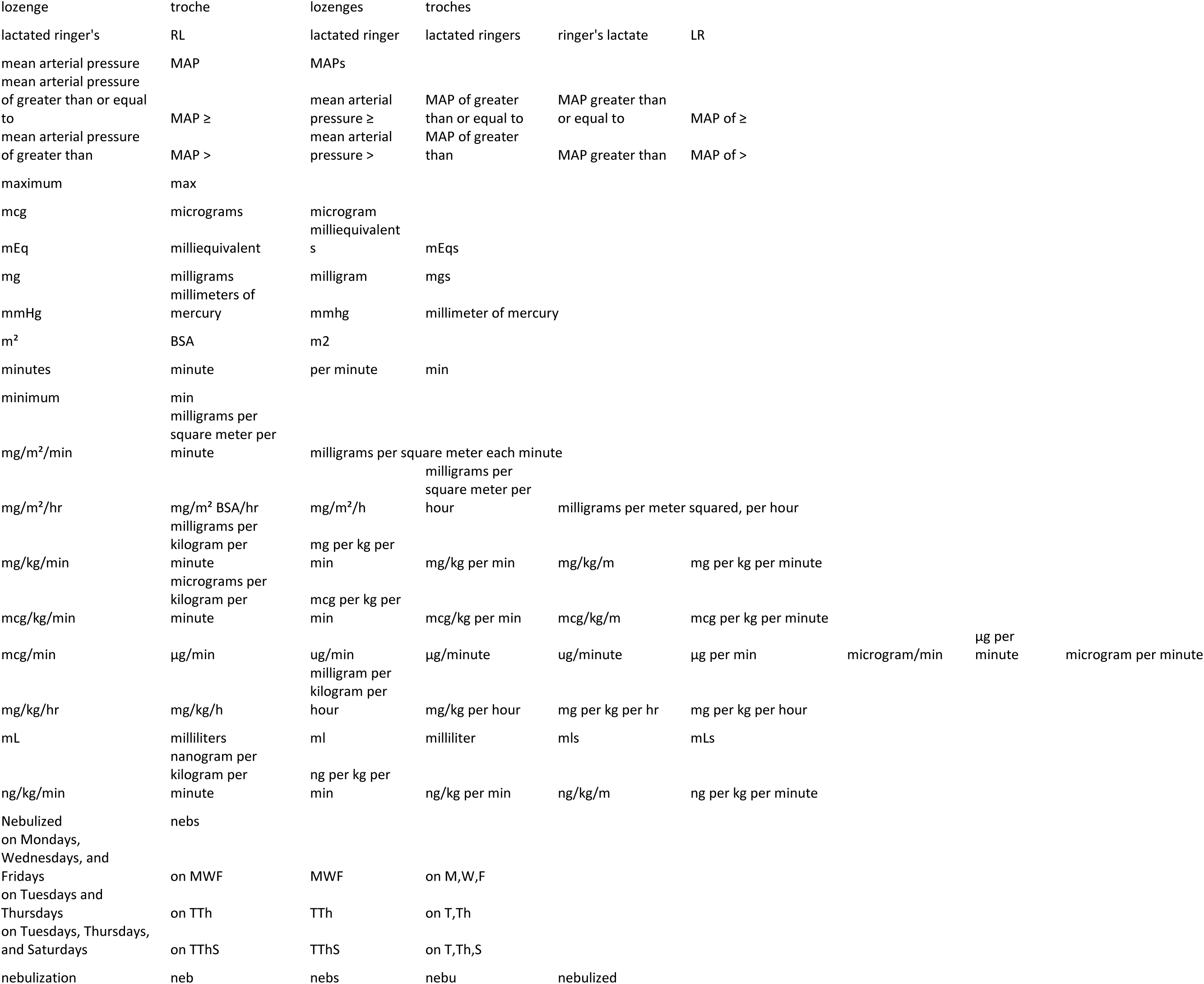
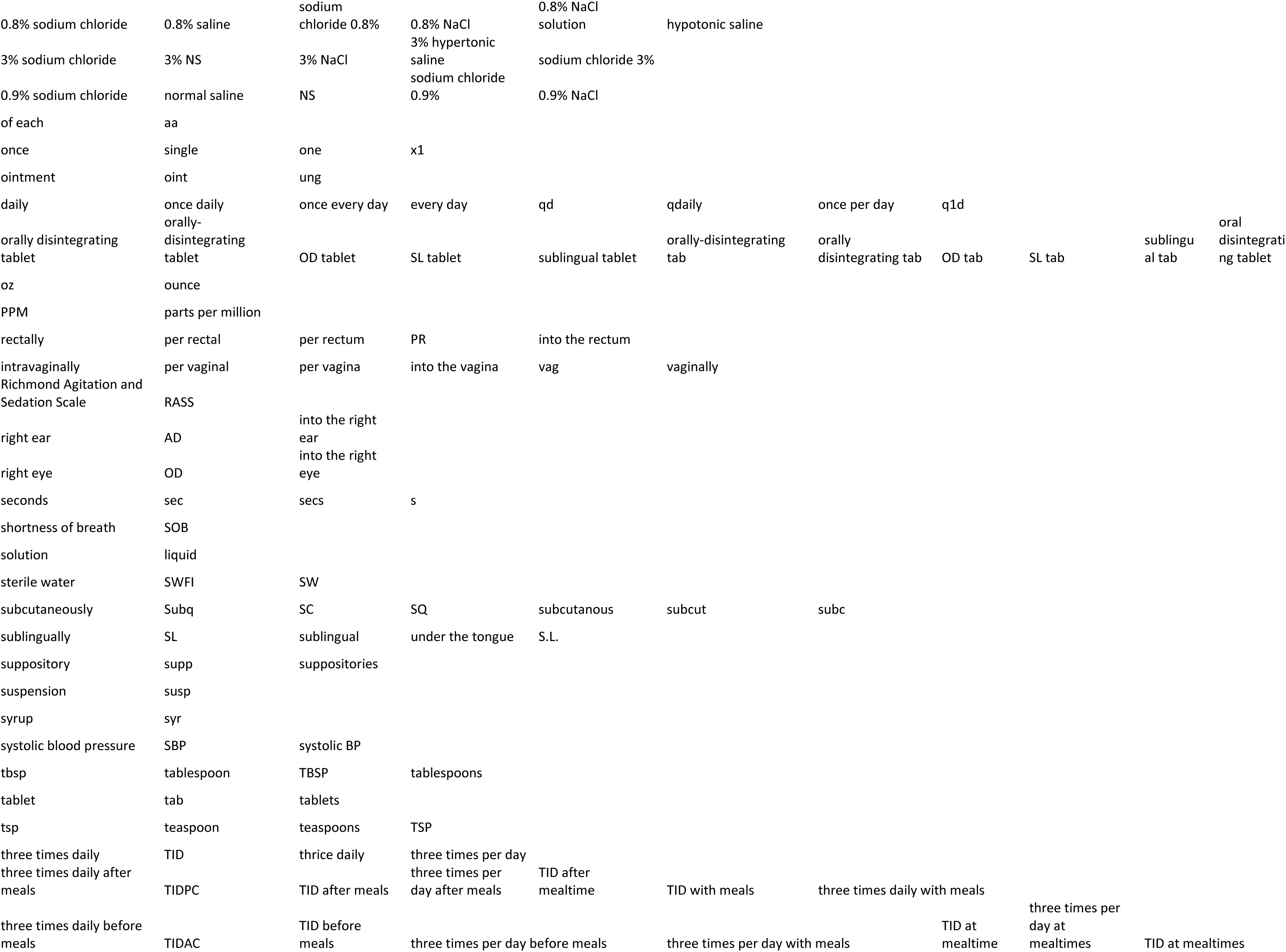
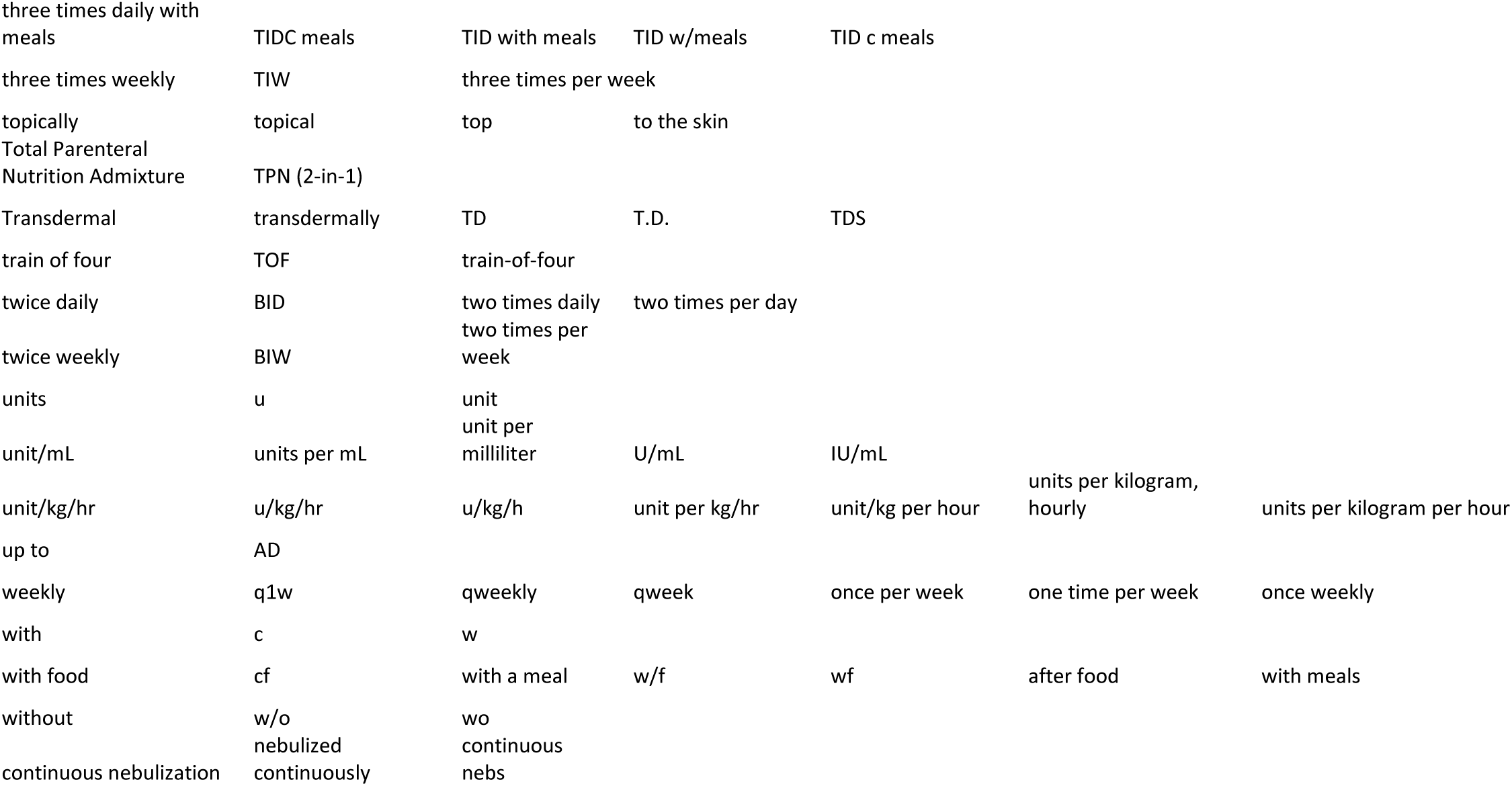

